# Validation of clinical diagnosis and machine learning classification of cognitive impairment

**DOI:** 10.64898/2026.07.21.26358593

**Authors:** Dan Mungas, Evan Fletcher, Keith Widaman, Sarah Tomaszewski Farias, Shraddha Sapkota, Pauline Maillard, Eleanor Hayes-Larson, L. Paloma Rojas-Saunero, Yixuan Zhou, Crystal Shaw, Elizabeth Rose Mayeda, Maria M. Corrada, Paola Gilsanz, Maria Glymour, John Olichney, Charles DeCarli, Rachel Whitmer, Brandon Gavett

## Abstract

**INTRODUCTION:** Cognitive syndrome diagnosis (Normal, Mild Cognitive Impairment (MCI), Dementia) is important for summarizing disease status and predicting future progression. Machine learning approaches to classification might substitute for or complement clinical diagnosis but must be shown to have validity for these purposes.

**METHODS:** A machine learning algorithm was trained in a previous study to reproduce clinical diagnosis of cognitive impairment [1]. We examined and compared concurrent validity (cross-sectional MRI measures of brain integrity) and predictive validity (longitudinal change in MRI measures of brain integrity and progression to a more impaired diagnosis/classification) of clinical diagnosis and algorithmic classification from the prior study.

**RESULTS:** Clinical diagnosis and algorithmic classifications had robust associations with clinical and MRI outcomes and differences across diagnosis/classification types were minor. Algorithmically estimated probability of a Normal diagnosis had the strongest associations with cross-sectional and longitudinal MRI outcomes. Progression from Normal to MCI or Dementia was faster for Clinical diagnosis than Algorithmic classification but future rates of MRI measured brain degeneration were essentially the same in individuals with baseline clinical diagnosis and algorithmic classification of Normal cognition.

**DISCUSSION:** Clinical diagnosis and algorithmic, machine learning based classification had robust and similar associations with independent validity criteria. Both forms of diagnosis/classification had utility for staging current brain degeneration and predicting future brain degeneration and clinical decline. This study demonstrates a validation design that can simultaneously evaluate the utility of both clinical diagnosis and algorithmic classification of cognitive impairment in a manner that improves understanding of both types of classification.

## 1. Introduction

Diagnosis is central to clinical health care – it summarizes complex clinical information to: a) identify presence of a disease or condition, b) identify an etiology when an abnormal condition is present, c) inform treatment/intervention decisions, and d) predict future disease progression.[2,3] Cognitive impairment and dementia in older adults are major public health problems and clinical diagnosis has a fundamental role in clinical care for those who are experiencing cognitive decline and dementia.[4] Delineation of cognitive and functional status is an important part of comprehensive clinical diagnosis.[4] This often is in the form of cognitive syndrome diagnosis (the focus of this study). Cognitive syndrome diagnosis typically summarizes stages of cognitive and functional impairment using three diagnostic categories: normal cognition, mild cognitive impairment (MCI), and dementia. These cognitive syndrome categories have differential practical implications for clinical treatment and care needs,[4,5] putatively represent increasing stages of brain degeneration,[6] and have differential value for predicting future brain degeneration [7] and rates of cognitive and functional decline.[8–10]

Cognitive syndrome diagnosis can be a resource intensive process, often requiring comprehensive clinical assessments and expert clinicians - resources that may be limited in clinical and research settings. Algorithmic classification might be useful as a proxy when clinical diagnosis is not available or feasible, and could provide a standardized and reproducible complement when clinical diagnosis is available.[1] Machine learning methods increasingly are being used to develop algorithms for syndrome classification [11]. The typical approach is to use expert clinician diagnosis as the standard and to develop models that combine predictive features to optimally reproduce clinician diagnosis. A variety of different machine learning methods and different sets of predictive features have been used in prior studies. Accuracy in predicting clinical diagnosis has been the main metric for evaluating performance of the machine learning models and has varied across studies from modest to excellent. The accuracy metric tells how closely the algorithm reproduces the clinical diagnosis, but it does not show how well the algorithmic classification serves important functions of diagnosis, notably disease staging and prognosis for future decline and impairment. Algorithmic classification that does not fully reproduce clinical diagnosis can still be valuable if it has staging and prognostic utility. Thus, comparing the concurrent and predictive validity of clinical diagnosis and algorithmic classification can show how effectively algorithmic classification can substitute for or complement clinical diagnosis. Such comparisons help to define the range of utility of both clinical and algorithmic classification methods.

This study is a follow-up to a previous study [1] that used a predictive feature set composed of measures of cognition and independent function along with demographic characteristics to predict clinical diagnosis in a demographically and clinically heterogenous sample. Accuracy of the algorithmic classification with respect to reproducing clinical diagnosis was moderate to strong but there were relevant differences in the classifications generated by these two approaches that suggest that algorithmic classification was not a direct substitute for clinical diagnosis. However, an important question remains about whether algorithmic classification in this setting can serve a similar function as clinical diagnosis.

In this study, we compared associations of clinical diagnosis and algorithmic classification with two sets of variables that were not used to derive either clinical diagnosis or algorithmic classification, cross-sectional and longitudinal MRI measures of brain integrity and future clinical progression to a more impaired syndrome diagnosis/classification. These MRI and clinical progression variables characterize disease staging and progression. This study examines how well algorithmic classification serves the staging and prognosis functions of clinical diagnosis. It also seeks to characterize the real world meaning and utility of both clinical diagnosis and algorithmic classification.

## 2. Methods

### 2.1. Overview of Study Design

This study built upon a previously developed predictive model [1] that used the XGBoost machine learning platform to predict clinical diagnosis in a merged sample of three longitudinal cohort studies of older adults: the UC Davis Alzheimer’s Disease Research Center Longitudinal Cohort (ADRC, N individuals = 1318, N assessments = 5445), the Kaiser Healthy Aging and Diverse Life Experiences Study (KHANDLE, N individuals = 628, N assessments = 1700), and the Life After 90 Study (LA90, N individuals = 824, N assessments = 3476. Features used to predict diagnosis included: high quality cognitive assessments, measures of independent function, demographic variables, and study cohort indicators. The predictive model developed in that study was then applied to all assessments for these three cohorts. This yielded predicted probabilities of Normal, MCI, and Dementia diagnoses and a predicted multiclass diagnosis (the diagnostic category with the highest probability) for every assessment in all three cohorts. All three cohorts had subsets of participants who had received research MRI scans following a standardized protocol. In the current study, clinical diagnosis and algorithmic classification were compared with respect to associations with two different types of criterion-related validation criteria: 1) rate of conversion from a baseline diagnosis/classification to a more impaired diagnosis/classification (predictive validity, all three cohorts), and 2) associations with cross-sectional (concurrent validity, all three cohorts) and longitudinal (predictive validity, ADRC only) quantitative MRI measures of brain integrity.

### 2.2. Sample Characteristics

The ADRC cohort includes substantial representation of Latino, Black, and non-Latino White older adults. Participants were recruited through 1) a community outreach and screening program designed to identify and recruit individuals with cognitive functioning representative of the community dwelling population in a six-county catchment area in the central Sacramento/San Joaquin valley and east San Francisco Bay area of Northern California, and 2) memory/dementia clinics [12]. This cohort has a rolling enrollment design in which new enrollees are added every year. Follow-ups occur on approximately an annual basis. This cohort was initiated in 2002 and individuals are followed until death or dropout. Participants in this cohort were assessed between 1 and 18 times for up to 21 years.

KHANDLE is comprised of community-dwelling older adults residing in the San Francisco Bay and Sacramento areas of California. Individuals eligible for KHANDLE were long-term members of Kaiser Permanente Northern California (KPNC), an integrated healthcare delivery system, were aged 65 years or older on January 1, 2017, spoke English or Spanish, and had previously participated in Kaiser Permanente multiphasic health checkup exams between 1964-1985. Individuals with a medical records diagnosis of dementia at time of enrollment were excluded. Stratified random sampling by race/ethnicity and educational attainment was used with the goal of recruiting approximately equal proportions of Asian, Black, Latino, and White participants and assuring diversity in educational attainment. A subset of KHANDLE participants received clinical assessments, including diagnosis, at each assessment wave. This clinical assessment subset included an approximately 30% random sub-sample, identified at study enrollment, who received a clinical evaluation at each wave plus individuals who screened positive for cognitive impairment at a specific assessment wave. At baseline in 2017, 1,712 individuals were enrolled in KHANDLE and a refresher cohort of ∼500 was added in 2022. This study included KHANDLE enrollees who had received a clinical diagnosis on at least one wave and included data from up to 4 waves with an average of about 1.4 years between assessments.

LA90 participants are long-term members of KPNC who were 90 years or older and residing in the San Francisco Bay and Sacramento areas of northern California. Eligible individuals were KPNC members at some point between 1964-1992 and spoke English or Spanish. A medical records diagnosis of dementia at the time of enrollment was an exclusion criterion. Non-White individuals were oversampled with a target recruitment goal 75% non-White. Study enrollment began in July 2018 and is ongoing, with 1322 enrolled through March, 2025. LA90 participants received clinical evaluations including cognitive syndrome diagnosis at all assessment waves, with follow-up at approximately 6 month intervals.

All participants in all three studies signed informed consent, and all human subject involvement was overseen by institutional review boards at UC Davis and Kaiser Permanente Northern California.

### 2.3. Clinical Evaluation and Diagnosis

#### 2.3.1. Clinical Evaluation Components

The clinical evaluation protocol for ADRC, KHANDLE, and LA90 included a clinical history and exam, administered by a trained clinician. Elements of the National Alzheimer’s Coordinating Center Uniform Dataset (NACC UDS)[13] were collected according to standardized UDS guidelines. ADRC and KHANDLE clinical evaluations included clinical neuropsychological assessment using the UDS Neuropsychological Battery.[14] The Modified Mini Mental State Exam (3MS)[15] was used to assess cognition in LA90 clinical evaluations. The examining clinician made a provisional clinical diagnosis of Normal cognition versus MCI versus Dementia for all three cohorts. The clinical evaluations protocol is described in detail in the prior publication.[1]

#### 2.3.2. Clinical Diagnosis

The same clinical diagnostic criteria were used for the ADRC, KHANDLE, and LA90. Dementia was diagnosed using DSM-III-R criteria for dementia modified such that dementia could be diagnosed in the absence of memory impairment if there was significant impairment of two or more cognitive domains. MCI was diagnosed according to standard clinical criteria according to UDS guidelines.[16] Normal cognitive function was diagnosed if there was no clinically significant cognitive impairment. Clinical diagnosis for the ADRC was made in a multidisciplinary case conference where all clinical evaluation findings incorporated in UDS instruments were available with the exception of the Clinical Dementia Rating (CDR;[17]), which was excluded from consideration in the case conference. UDS neuropsychological test results were considered. KHANDLE and LA90 diagnoses were derived by applying standardized, rational-intuitive decision rules to the data elements collected during the clinical evaluation. This was designed to recapitulate the provisional clinical syndrome diagnostic criteria used by clinicians and is described in detail in the previous publication [1].

### 2.4. Predictive Model Development

All three studies included measures of cognition (Spanish and English Neuropsychological Assessment Scales (SENAS)[18,19]) and independent function (Everyday Cognition (ECog)[20]) that were administered at every assessment wave but were not used for clinical diagnosis. These measures, demographic characteristics (age, level of education, sex/gender, and race/ethnicity) and indicator variables for cohort were used as predictive features to train the XGBoost model to predict diagnosis in ADRC, KHANDLE, and LA90. These measures were available at all assessments for these three cohorts. This made it possible to generate a predicted diagnosis at every assessment for the three cohorts. A 10-fold cross-validation design was used such that results of a model trained on 9 folds could be used to generate out-of-sample predictions in the 10th, held out fold. This was repeated 10 times to generate out-of-sample predictions for all assessments involving the three cohorts. Detailed information about the model training and out-of-sample prediction is available in the previous publication [1].

### 2.5 MRI Methods

MRI brain image acquisition was performed at the UC Davis Imaging Research Center (IRC) or at the Veterans Administration Northern California Health System Medical Center in Martinez, CA (Martinez VA) on 1.5 and 3T MRI scanners using a standardized image acquisition protocol developed and overseen by the Imaging Core of the UC Davis Alzheimer’s Disease Research Center. All images were processed in-house at the IDeA Laboratory, Department of Neurology, University of California, Davis, as described in previous publications.[21–23]

MRI measures of brain integrity, selected to capture complementary aspects of brain integrity relevant to cognitive impairment, included: 1) total gray matter volume, 2) hippocampal volume, 3) union signature, and 4) cingulum-hippocampus tract free water. Total gray matter and hippocampal volumes are quantitative measures of segmented gray matter in these regions of interest, and the processing pipeline to derive these variables has been previously described (e.g. [22,24,25]). Union signature is a computationally derived “signature” of cortical gray matter thickness in regions of interest that optimally explain episodic memory performance. It was originally developed with the Alzheimer’s Disease Neuroimaging Initiative (ADNI) sample and data but has since been cross-validated with ADRC, KHANDLE, and LA90.[26,27] It is strongly associated with several clinical/cognitive outcomes including episodic memory, executive function, CDR Sum, and clinical syndrome diagnosis, outperforming associations by known gray matter regions such as the hippocampus and medial temporal ROIs and previously published Alzheimer’s disease signatures.[28] Free water measures of white matter tracts are computed by modeling diffusion tensor images (DTI) as two compartments: a free water compartment, representing extracellular water molecules, and a tissue compartment, accounting for all other molecules [29]. A larger fraction for the free water compartment indicates degraded cellular and axonal structure. Thus, lower free water values represent greater white matter tract integrity. Free water has been shown to be a sensitive marker of cognition and clinical status [23]. Free water in the part of the cingulum bundle connecting to the hippocampus via the parahippocampal gyrus (cingulum-hippocampus) was selected for this study because it has the strongest associations with cognitive outcomes in the three study cohorts.

Union signature, total gray matter volume, and hippocampal volume were harmonized to minimize scanner effects – unwanted and non-biological site-specific additive and multiplicative bias – due to scanner field strength. The freewater cingulum-hippocampus measure was harmonized based on field strength and the number DTI vector directions. We used the ComBat (“combating batch effects when combining batches”) method [30] to adjust for scanner (“batch”) effects on MRI variables. This method has been used to harmonize both structural MRI and DTI [31] and we used a recent adaption for longitudinal data developed by Beer [32] that is implemented in the R longCombat package (version 0.0.0.90000). This harmonization effectively removed non-biological bias associated with scanner field strength and number of directions in the DTI vector. Field strength associations with MRI variables, the approach to field strength adjustment of MRI measures, and comparisons of adjusted and unadjusted MRI variables are described in detail in Appendix A.1.1. Supplemental Table 1 shows detailed results of effects of biological variables (age, sex), conceptually relevant covariates (baseline Clinical diagnosis, time-varying Algorithmic probability of a Normal diagnosis), time-from-1st-scan, scanner field strength, and number of DTI directions on pre-harmonization and post-harmonization MRI variables. MRI variables were Blom standardized [33] to facilitate interpretability of results, and this is described in Appendix A.1.1.

### 2.6 Data Analysis

#### 2.6.1 Diagnostic Conversion

Time to event analyses were performed to compare clinical and algorithmic diagnoses with respect to rate of progression to a more impaired cognitive syndrome diagnosis. We examined three specific types of diagnostic progression: 1) Conversion from Normal cognition to cognitive impairment (MCI or Dementia), 2) Conversion from Normal cognition to Dementia, and 3) Conversion from MCI to Dementia. Algorithmic classification for these categories was derived from the cross-validated, multiclass predicted diagnosis generated by the XGBoost prediction model. The most probable class was selected. For example, MCI would be the classification if the probability of MCI exceeded the probability of Normal and the probability of Dementia. ADRC, KHANDLE, and LA90 participants were used for these analyses. For each type of conversion, a dataset was created by selecting individuals who had the “conversion-from” classification at the baseline evaluation (e.g. Normal) and had three or more longitudinal evaluations for which a clinical and an algorithmic classification was available. This dataset was converted to a time-to-event format with time from the baseline assessment as the time variable, diagnosis as the outcome (1 if conversion-to classification, 0 otherwise), and a censored indicator variable (1 if lost to follow up or deceased before a conversion occurred or still being followed without a conversion, 0 otherwise). Importantly, each dataset included only individuals who had both the conversion-from clinical diagnosis and the conversion-from algorithmic classification at the baseline assessment. This established a common sample for comparing the two diagnosis/classification types. In the MCI conversion to dementia analysis, for example, the sample was composed of those with both a clinical and algorithmic classification of MCI. Consequently, the follow-up sample was identical for both types of diagnosis.

The R survival package (version 3.6-4) was used to estimate accelerated failure time (AFT) survival models and to compare rates of survival without conversion for the three specific dichotomous diagnoses across clinical and algorithmic classification types. AFT survival models are parametric models, with flexible options for the baseline distribution function. We fitted the same models with multiple different baseline distributions. The lognormal distribution consistently resulted in the best model fit and was used in subsequent analyses.

#### 2.6.2. Diagnosis Association with MRI Measures of Brain Structure - Cross-Sectional

We performed a series of analyses to examine cross-sectional associations of MRI measures of brain integrity with diagnosis/classification variables and compare these results across diagnosis/classification variable types. Three different types of diagnosis/classification variables were compared (clinical - multiclass, algorithmic - multiclass, algorithmic - continuous). All three cohorts (ARDC, KHANDLE, LA90) were included in these analyses. Cross-sectional associations of different types of diagnosis/classification with MRI measures were evaluated with bivariate linear regression models in which the MRI variables were dependent variables and diagnosis/classification category was the independent variable. The primary statistic of interest was the *R*^2^ value, summarizing the amount of variance in the MRI variable explained by the diagnosis/classification. Higher values indicate a stronger association. Root mean squared error (RMSE) was a supplemental indicator of model fit. RMSE shows the predictive accuracy of the model with lower values indicating better fit. These two model fit statistics allowed for direct comparison of associations of multiclass and continuous probability of diagnosis/classification variables with MRI variables. The datasets for each of these analyses included the MRI measure from the first obtained scan and the diagnosis/classification variables from the clinical evaluation closest in time to that scan (and within 1.5 years from the scan). There was an additional requirement that both clinical and algorithmic diagnosis/classification were available so that the analyses for both were based on the same sample. Bootstrap confidence intervals for *R*^2^ and RMSE estimates were created by using sampling with replacement to generate 10,000 bootstrap samples and estimate the *R*^2^ and RMSE statistics in each sample. These bootstrap samples were also used to generate 95% confidence intervals for differences between two different diagnosis/classification variables in *R*^2^ or RMSE.

#### 2.6.3. Diagnosis Association with MRI Measures of Brain Structure - Longitudinal

##### 2.6.3.1. Overview

Analyses of longitudinal MRI variables in relation to diagnosis/classification type variables were restricted to the ADRC sample because only 1 scan was available for LA90 and KHANDLE participants mostly had 1 scan (388 out of 435 participants with MRI). Mixed effects longitudinal models were estimated - Blom standardized MRI variables were dependent variables and diagnosis/classification variables were independent variables. An intercept random effect was estimated in each model. This uses all MRI observations for an individual to estimate a person-specific value at the time of the first scan that accounts for within-person random error. A time-from-1st-scan fixed effect was included in all models to evaluate the average annual change in the MRI variable per 1 year of follow-up. Two different types of analysis were performed. In the first, diagnosis/classification was added to time-from-1st-scan as a time-varying fixed effect in the model. In the second type of analysis, baseline diagnosis (at the time of the first scan) was the diagnosis variable. This model included three fixed effects: time-from-1st-scan, diagnosis/classification (effect of baseline diagnosis/classification on estimated MRI variable intercept), and time-from-1st-scan by diagnosis/classification interaction. The R lme4 package (version 1.1-35.5) was used to estimate mixed effects models. R multcomp (version 1.4-26)::glht() was used to estimate and evaluate statistical significance of linear combinations and contrasts of effects from the mixed effects models. This generates standard errors and confidence intervals for different combinations of effects in the model, and for example, allows for estimating the difference between effects of two different diagnosis/classification types on a MRI variable, with an associated standard error that can be used for significance testing. It is important to note that corrections for multiple comparisons were not applied to statistical significance tests associated with reported comparisons. An implication of this decision is that it will be more likely to reject the null hypothesis that effects of clinical and algorithmic classification do not differ. In addition, model performance was compared across different types of diagnosis/classification in the different longitudinal analysis designs. R performance (version 0.12.3) was used to compare models using marginal *R*^2^ as the primary model fit metric. Marginal *R*^2^ represents the amount of total variance in the mixed effects model that is explained by the fixed effects. In this study, this corresponds to the total variance in the MRI variable that was explained by the fixed effects included in the different models.

##### 2.6.3.2. Time-varying Diagnosis

MRI variables were dependent variables in linear mixed effects models that included a person random effect and time and time-varying diagnosis as fixed effect independent variables. The general symbolic formula for these analyses was:

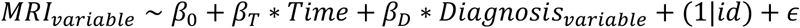

where *MRI_variable_* is the MRI variable, Time = time from 1st scan, *Diagnosis_variable_* is the time-varying diagnosis/classification variable, *β*_0_, *β_T_*, and *β_D_* are fixed effects regression coefficients, (1|id) specifies a person (intercept) random effect, and *ε* is random error. The *β_D_* coefficient captures average differences across diagnosis categories and combines between-person differences at baseline with effects of within person change in diagnosis over time. Separate analyses were performed for each combination of MRI variables (union signature, freewater cingulum-hippocampus, hippocampus, total gray matter) and three diagnosis/classification variables (multiclass Clinical, multiclass Algorithmic, and continuous probability of Normal diagnosis).

##### 2.6.3.3. Baseline Diagnosis

Baseline diagnosis variables were used in these analyses as independent variables to explain MRI variable intercepts and as modifiers of the Time fixed effect. The models did not include within-person diagnosis change effects on MRI variables. The general symbolic formula was:

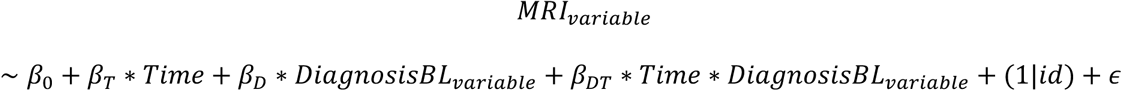

*β_D_* captures the effect of baseline diagnosis/classification on the MRI variable intercept (i.e., at baseline) and *β_DT_* is an interaction effect that captures how the average Time effect differs across baseline diagnosis groups.

#### 2.6.4. Longitudinal Change in Probability of Dementia in Relation to MRI Variable at Baseline and MRI Variable Change

A separate set of analyses examined how repeated-measures diagnosis/classification variables were associated with MRI independent variables. These analyses were restricted to the ADRC cohort and evaluated how probability of dementia changed over time as a function of a baseline brain integrity and longitudinal change in brain integrity. Diagnosis variables for these analyses were: 1) dichotomous clinical diagnosis of dementia, 2) dichotomous algorithmic classification of dementia, and 3) continuous algorithmically predicted probability of dementia. These longitudinal diagnosis/classification outcomes were modeled in generalized linear mixed effects models using a baseline MRI variable and change in that MRI variable as independent variable predictors of diagnosis/classification. Analyses for the two dichotomous variables used a Bernoulli family distribution with a logistic link. Beta family with logistic link was used for continuous probability of dementia. This expresses all three outcomes on a 0-1 probability of dementia scale. As in previous analyses, MRI variables were field strength, DTI vector dimension, and intracranial volume adjusted and Blom standardized. Baseline MRI values were the standardized values from the first scan and change for an individual was calculated as a standardized value for a scan minus the standardized value for the first scan (change for the first scan was 0).

The general symbolic formula for these analyses was:

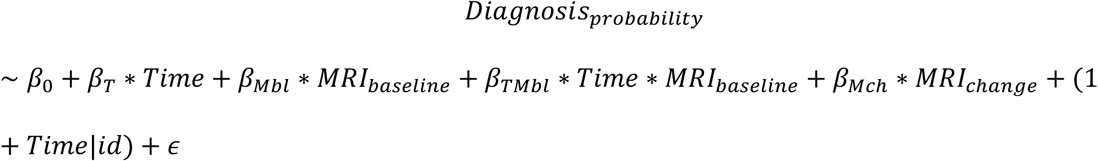

where *Diagnosis_probability_* is probability of Dementia predicted by the diagnosis variable, Time = time from 1st scan, *MRI_baseline_* is the MRI variable value for the baseline scan, *MRI_change_* is the MRI variable value minus the value for the baseline scan, *β*_)_, *β_T_*, and *β_Mbl_*, *β_Mc_*_h_, and *β_TMbl_* are fixed effects regression coefficients, (1+Time|id) specifies person-specific intercept and slope random effects, and *ε* is random error. There were sufficient longitudinal diagnoses in the ADRC sample to enable estimation of slope random effects.

Bayesian regression was used for model estimation implemented by R brms (version 2.22.0). brms has an option for imputing missing values of variables within an analysis using model-derived estimates. For these analyses, the number of scans was smaller than the number of clinical evaluations and the brms mi() option was used to replace missing *MRI_change_* values using the following model:

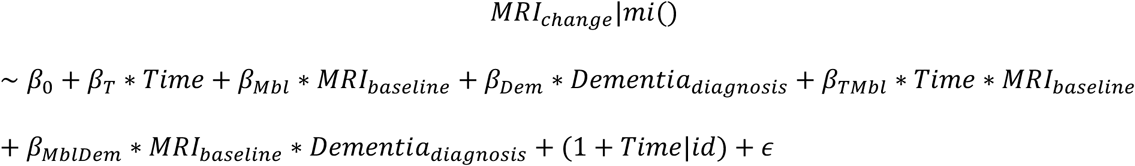

where *MRI_change_*|*mi*() indicates that missing *MRI_change_* values for assessments will be replaced by imputed data using a formula in which observed *MRI_change_* is predicted by Time from the baseline assessment, *MRI_baseline_*, *Dementia_diagnosis_* at the missing assessment (time-varying), a Time X *MRI_baseline_* interaction, and a *MRI_baseline_* X *Dementia_diagnosis_* interaction; *β*_)_, *β_T_*, *β_Mbl_*, *β_Dem_*, *β_TMbl_*, and *β_MblDem_* are fixed effects regression coefficients, (1+Time|id) specifies person-specific intercept and slope random effects, and *ε* is random error.

## 3. Results

### 3.1. Sample Characteristics

Table 1 shows sample characteristics by study cohort. Baseline prevalence of clinical diagnosis of Normal cognition was lowest in ADRC (58%) and was higher and similar in KHANDLE (76.5%) and LA90 (79.5%). MCI clinical diagnosis baseline prevalence was 26.2% in the combined sample and varied across study cohorts from 19.7% (LA90) to 30.1% (ADRC).

**Table 1.** Sample characteristics by study cohort. Column totals are numbers of individuals in each cohort.

|  | ADRC<br>(N=1153) | KHANDLE<br>(N=409) | LA90<br>(N=488) | Overall<br>(N=2050) |
| --- | --- | --- | --- | --- |
| **diagnosis (baseline)** |  |  |  |  |
| Normal | 660 (58.0%) | 313 (76.5%) | 388 (79.5%) | 1361 (66.9%) |
| MCI | 342 (30.1%) | 94 (23.0%) | 96 (19.7%) | 532 (26.2%) |
| Dementia | 135 (11.9%) | 2 (0.5%) | 4 (0.8%) | 141 (6.9%) |
| **sex/gender** |  |  |  |  |
| Mean (SD) | 1.61 (0.487) | 1.56 (0.497) | 1.61 (0.488) | 1.60 (0.490) |
| Median [Min, Max] | 2.00 [1.00, 2.00] | 2.00 [1.00, 2.00] | 2.00 [1.00, 2.00] | 2.00 [1.00, 2.00] |
| **age (baseline) (years)** |  |  |  |  |
| Mean (SD) | 75.0 (7.28) | 75.9 (6.50) | 92.1 (2.09) | 79.2 (9.55) |
| Median [Min, Max] | 75.0 [49.0, 97.0] | 75.2 [65.6, 99.0] | 91.4 [90.1, 103] | 78.0 [49.0, 103] |
| **education (years)** |  |  |  |  |
| Mean (SD) | 13.5 (4.56) | 14.8 (2.99) | 14.3 (3.19) | 13.9 (4.03) |
| Median [Min, Max] | 14.0 [0, 20.0] | 16.0 [2.00, 20.0] | 14.0 [0, 20.0] | 14.0 [0, 20.0] |
| **race** |  |  |  |  |
| Asian | 48 (4.2%) | 96 (23.9%) | 119 (25.1%) | 263 (13.0%) |
| Black | 286 (24.9%) | 80 (20.0%) | 113 (23.8%) | 479 (23.7%) |
| LatinX | 281 (24.5%) | 112 (27.9%) | 89 (18.7%) | 482 (23.8%) |
| NativeAmer | 4 (0.3%) | 0 (0%) | 0 (0%) | 4 (0.2%) |
| White | 528 (46.0%) | 113 (28.2%) | 150 (31.6%) | 791 (39.1%) |
| Native American | 0 (0%) | 0 (0%) | 4 (0.8%) | 4 (0.2%) |
| **N assessments** |  |  |  |  |
| Mean (SD) | 4.44 (3.51) | 3.12 (1.03) | 4.78 (2.04) | 4.26 (2.91) |
| Median [Min, Max] | 4.00 [1.00, 18.0] | 3.00 [1.00, 4.00] | 4.00 [1.00, 8.00] | 4.00 [1.00, 18.0] |
| **fu_time (years)** |  |  |  |  |
| Mean (SD) | 4.53 (4.48) | 4.62 (1.52) | 2.38 (1.13) | 4.04 (3.59) |
| Median [Min, Max] | 3.45 [0, 20.5] | 5.14 [0, 6.64] | 2.30 [0, 4.31] | 3.43 [0, 20.5] |
| **diagnosis (baseline) -**<br>**MRI (cross-sectional)** |  |  |  |  |
| Normal | 631 (57.6%) | 221 (78.4%) | 160 (78.8%) | 1012 (64.1%) |
| MCI | 329 (30.0%) | 59 (20.9%) | 39 (19.2%) | 427 (27.0%) |
| Dementia | 135 (12.3%) | 2 (0.7%) | 4 (2.0%) | 141 (8.9%) |
| **diagnosis (baseline) -**<br>**MRI (longitudinal)** |  |  |  |  |
| Normal | 351 (66.5%) |  |  | 351 (66.5%) |
| MCI | 147 (27.8%) |  |  | 147 (27.8%) |
| Dementia | 30 (5.7%) |  |  | 30 (5.7%) |
| **N MRI (longitudinal)** |  |  |  |  |
| 2 | 290 (54.9%) |  |  | 290 (54.9%) |
| 3 | 136 (25.8%) |  |  | 136 (25.8%) |
| 4+ | 102 (19.3%) |  |  | 102 (19.3%) |

Dementia clinical diagnosis prevalence was 11.9% in ADRC but was about 1% in KHANDLE and LA90, where a medical records diagnosis of dementia excluded enrollment. Gender distribution was similar across cohorts. Baseline age was much greater in LA90, as would be expected, and was similar in KHANDLE and ADRC. Average education ranged from 13.5 years in ADRC to 14.8 in KHANDLE, with LA90 intermediate at 14.3. KHANDLE and LA90 had relatively good balance across the 4 main race/ethnicity groups, whereas ADRC included proportionally more White (46% of sample) and relatively few Asian (4.2% of sample) participants. Average number of assessments was highest in LA90 (4.8), lowest in KHANDLE (3.1), and intermediate in ADRC (4.4). Average time of follow-up was about the same in in ADRC (4.5 years) and KHANDLE (4.6 years) and was lowest in LA90 (2.4 years) due to competing risk of death in this aged cohort. The ADRC sample included 462 individuals who had 5 or more assessments, LA90 had 226, and the maximum number of assessments in KHANDLE was 4.

Cross-sectional MRI results were available for 1580 participants (N ADRC = 1095, N KHANDLE = 282, N LA90 = 203). Prevalence of Dementia (clinical diagnosis) at the time of the scan was higher in ADRC (12.3%) than in KHANDLE (0.7%) and LA90 (2%), and conversely, prevalence of Normal cognition (clinical diagnosis) was lower in ADRC (57.6%) than in KHANDLE (78.4%) and LA90 (78.8%). Longitudinal MRI was available for 528 ADRC participants. The prevalence of Dementia at the baseline scan was lower (5.7% vs. 12.3%) and prevalence of Normal was higher (66.5% vs. 57.6%) compared to the ADRC cross-sectional MRI sample. A majority of participants (54.9%) had received 2 scans and 45.1% had received 3 or more.

Participants from the overall sample described in Table 1 were involved in one or more of the analyses for this study, all of which had specific inclusion criteria. Supplemental Table 2 presents sample characteristics for those included in the different types of analyses. For diagnosis conversion analyses, 3+ assessments with clinical and algorithmic classification were required and individuals from all three cohorts were included. One set of analyses examined progression from a baseline classification of Normal to Impaired (MCI or Dementia, N=1009), a second from Normal to Dementia (N=1009) and a third from MCI to Dementia (N=249). Cross-sectional MRI analyses included individuals from all three cohorts (N=1580) - longitudinal MRI analyses were restricted to the ADRC cohort (N=528 individuals, 1438 total MRI scans).

Distributions of sex/gender were similar across the four analysis types presented in Supplemental Table 2 (% female ranging from 55.4-63.8%). Baseline age was 3-8 years younger on average in the MRI analysis subsamples. Average education was about 14 years in three of the analysis subsamples and was 14.6 in the Normal to MCI/Dementia conversion subsample.

Distribution of race/ethnicity was similar across the four subsamples with the exception of the longitudinal MRI subsample; this study was restricted to the ADRC and consequently Whites were more strongly represented and Asians less represented. Number of assessments and follow-up time were greater in the longitudinal MRI subsample and the Normal to MCI/Dementia conversion subsample.

### 3.2. Diagnosis/Classification Conversion

#### 3.2.1. Normal to Impaired (MCI or Dementia)

Diagnosis/classification conversion was examined in a sample that included ADRC, KHANDLE and LA90 cohorts. There were 1009 individuals at risk for conversion from Normal to Impaired (MCI or Dementia); 362 converted in 4254 person-years for Clinical diagnosis and 241 for Algorithmic classification. Impairment free survival time significantly differed across diagnosis types (classification type = Clinical, reference type = Algorithmic, coefficient = -0.41, SE = 0.07, z = -6.08, p = 0, exp(coefficient) = 0.664). Impairment-free time to 50% survival was about 33.6% shorter for Clinical diagnosis (6.85 years) compared to Algorithmic classification (10.29 years). The left pane of Figure 1 shows the faster conversion time for Clinical diagnosis than for Algorithmic classification. A secondary analysis examined conversion from Normal to MCI. Results were similar to those involving conversion from Normal to Impaired with 37.5% faster progression to MCI for Clinical diagnosis (classification type = Clinical, reference type = Algorithmic, coefficient = -0.47, SE = 0.07, z = -6.53, p = 0, exp(coefficient) = 0.625).

**Figure 1.**
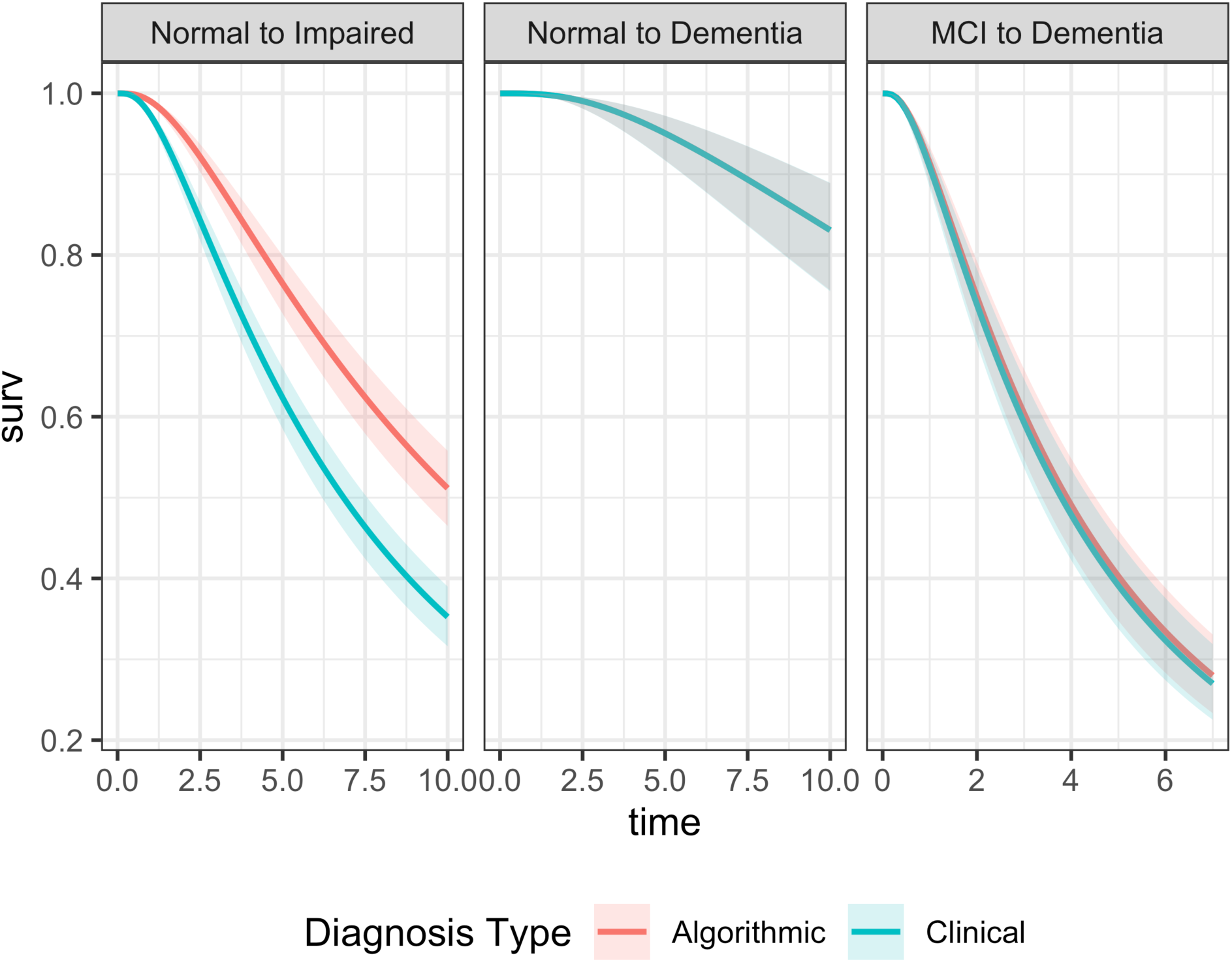
Accelerated failure time survival curves by diagnosis type and type of conversion. [Note. Survival curves for Normal to Dementia diagnosis progression are overlapping and appear as one curve.]

#### 3.2.2. Normal to Dementia

There were 1009 individuals at risk for conversion from Normal to Dementia; 62 converted in 4404 person-years for clinical diagnosis and 66 converted for algorithmic classification. Dementia-free survival time did not differ by diagnosis type (classification type = Clinical, reference type = Algorithmic, coefficient = 0, SE = 0.1, z = 0, p = 1, exp(coefficient) = 1). The AFT predicted survival curves (Figure 1, middle pane) are virtually the same.

#### 3.2.3. MCI to Dementia

There were 249 individuals at risk for conversion from MCI to Dementia - there were 122 conversions for clinical diagnosis and 112 for algorithmic classification in 830 person-years follow-up time. Dementia-free survival time did not differ by diagnosis type (classification type = Clinical, reference type = Algorithmic, coefficient = -0.03, SE = 0.1, z = -0.31, p = 0.76, exp(coefficient) = 0.97) and there was minimal divergence of the AFT predicted survival curves (Figure 1, right pane).

#### 3.2.4. Summary - Diagnosis Conversion by Diagnosis Type

Clinical diagnosis progressed faster than algorithmic classification from Normal to Impaired but rate of progression to Dementia, from either Normal or MCI, did not differ by diagnosis type.

### 3.3. MRI and Diagnosis Association

#### 3.3.1. MRI and Diagnosis Association - Cross-sectional

Cross-sectional MRI-Diagnosis associations were examined in the combined sample of all three cohorts. Figure 2 shows *R*^2^ and RMSE values for all of the combinations of MRI and diagnosis variables. Algorithmically estimated probability of being Normal (magenta) had the strongest association with all four MRI variables (highest *R*^2^) and best model fit (lowest RMSE). Statistical significance of differences in *R*^2^ and RMSE values for two different diagnosis/classification variables was evaluated by determining if the 95% bootstrap confidence interval for the difference did not include 0. The *R*^2^ and RMSE values for probability of Normal were significantly different from those statistics for the four other diagnosis/classification variables for all four MRI variables. Clinical diagnosis and Algorithmic classifications did not significantly differ. Continuous probability of Dementia (green) was equal to or slightly poorer than the two multiclass diagnoses and probability of MCI consistently had the weakest association/model fit. Supplemental Table 3 provides a tabular version of the results presented in Figure 2.

**Figure 2.**
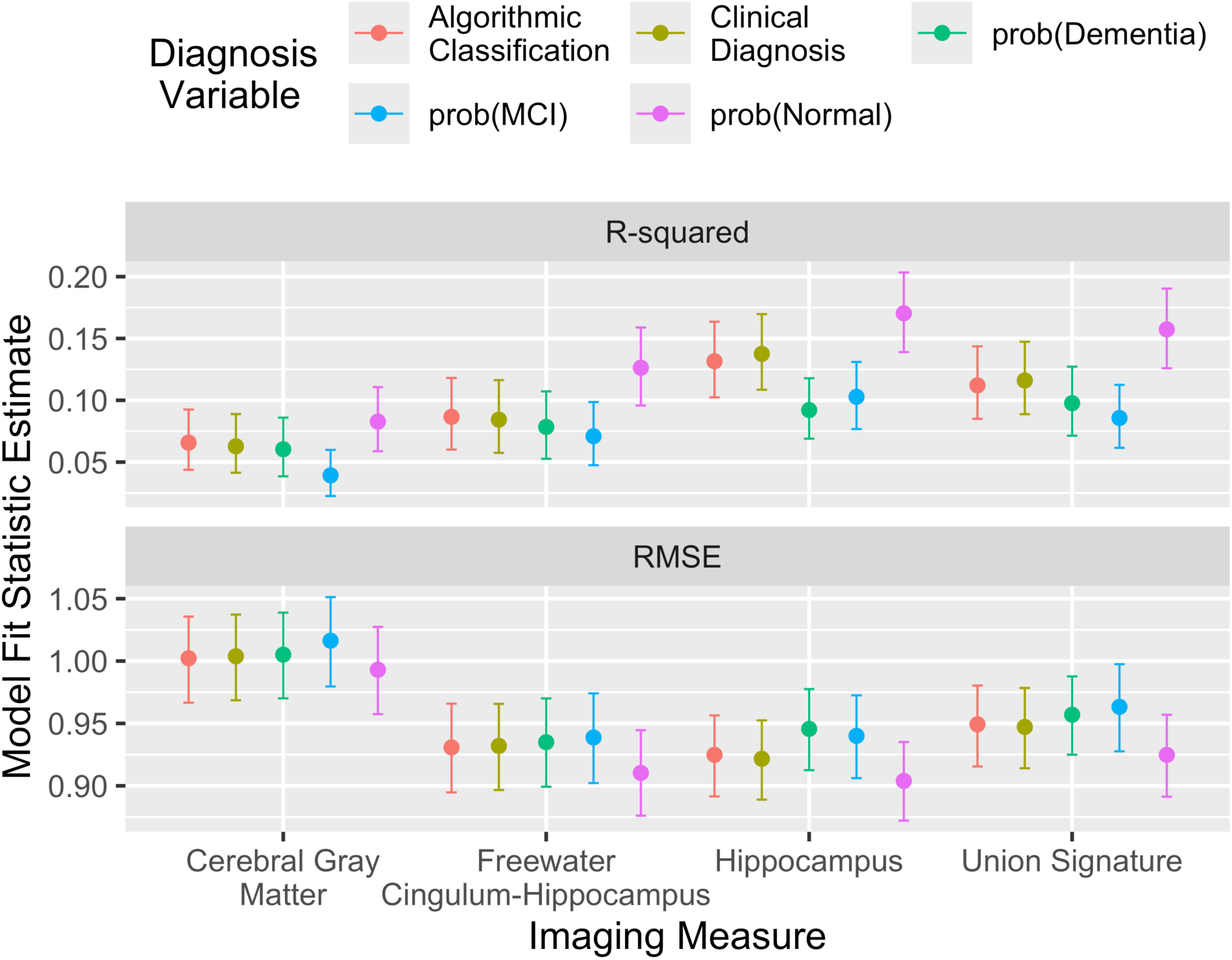
Model fit of MRI brain measures predicted by diagnosis variables - all studies, cross-sectional. [Dots show average across 10,000 bootstrap sample and bars show 95% confidence intervals.]

#### 3.3.2. MRI and Diagnosis Association - Longitudinal

Longitudinal analyses involving MRI were restricted to the ADRC sample. Time varying diagnosis effects combine average between-person differences across diagnostic categories with average within person differences associated with within-person change in diagnosis. The pattern of results for summary indices was quite similar to that for cross-sectional MRI-diagnosis analyses. Marginal *R*^2^ values were highest for continuous probability of a Normal diagnosis and were lower and very similar across categorical Clinical and Algorithmic diagnosis types (see Supplemental Table 4). For example, marginal *R*^2^ for Union Signature was 0.243 for continuous probability of Normal, 0.176 for Clinical categorical diagnosis, and 0.196 for Algorithmic classification.

Figure 3 summarizes time-varying diagnosis effects on MRI brain variables for Clinical diagnosis and Algorithmic classification. There were clearly apparent differences across diagnosis classes/categories (Normal, MCI, Dementia), but results within each class did not differ across diagnosis/classification types. Supplemental Table 5 presents a tabular version of results in Figure 3, and in addition, shows model estimated Clinical-Algorithmic differences for each diagnosis/classification category (Normal, MCI, Dementia).

**Figure 3.**
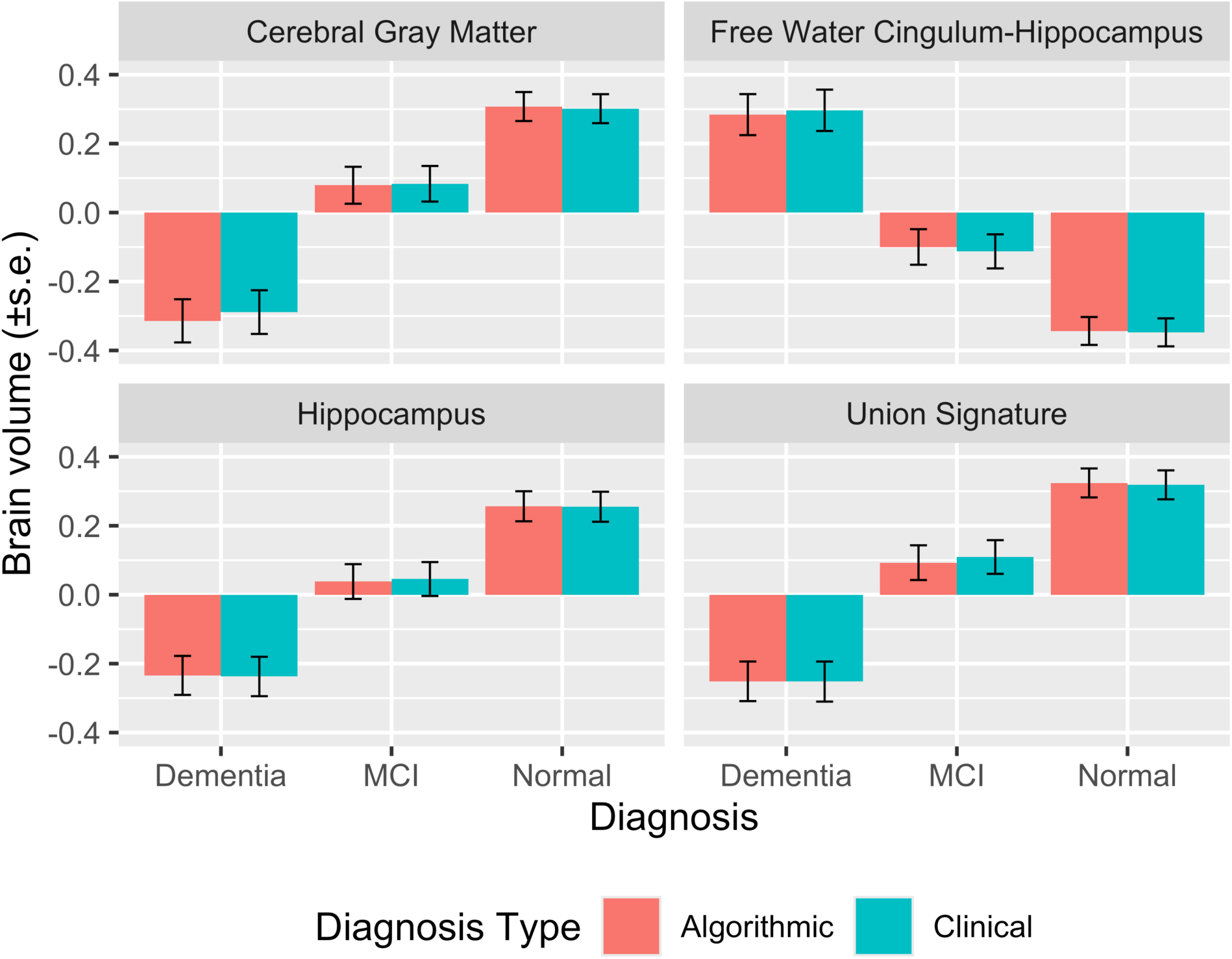
Plot of time-varying diagnosis effect on brain measures by diagnosis type - longitudinal analysis (ADRC Cohort).

Analyses examining effects on MRI variables of baseline diagnosis and its interaction with Time also showed that continuous probability of Normal had the strongest association with longitudinal MRI variables while the categorical Clinical diagnosis and Algorithmic classification variables had lower and similar marginal *R*^2^ values (see Supplemental Table 6).

Figure 4 is a plot of the effects of baseline diagnosis/classification on MRI variable intercepts derived from results of the mixed effects longitudinal model. The diagnosis category differences are less pronounced than for the time-varying diagnosis effects in Figure 3, but again, results within each category do not significantly differ across diagnosis types.

**Figure 4.**
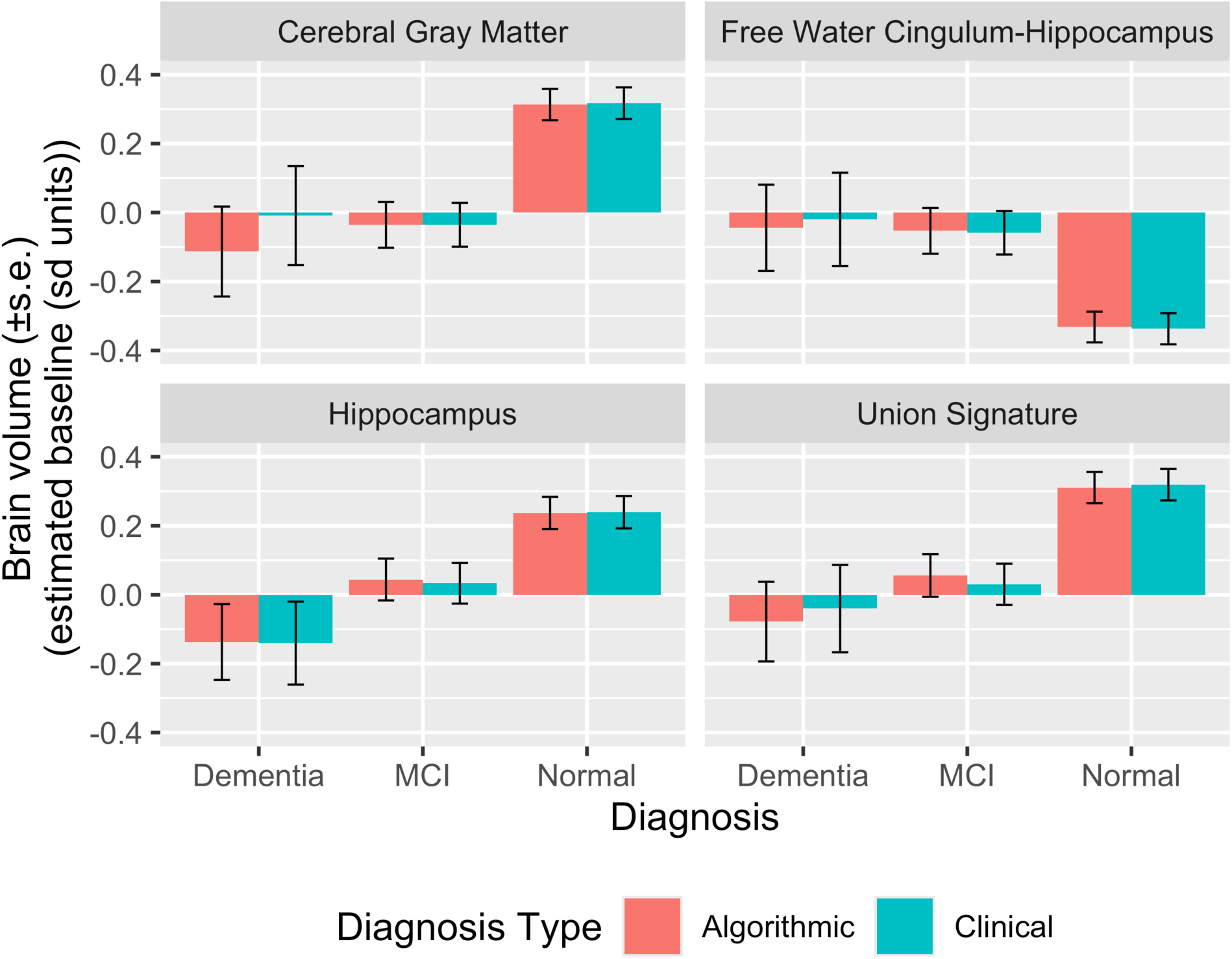
Plot of baseline diagnosis effects on brain measure intercepts by diagnosis type - longitudinal analysis (ADRC Cohort).

Comprehensive statistical significance tests for these analyses are presented in Supplemental Table 7.

Effects of baseline diagnosis/classification on MRI variable change derived from the mixed effects longitudinal models are presented in Figure 5. This figure shows the average annual change of MRI variables (in SD units) across diagnosis/classification categories for clinical diagnosis and algorithmic classification. Again it is most noteworthy that the effects of baseline diagnosis on MRI variable change do not appreciably differ within diagnostic/classification category across Clinical diagnosis and Algorithmic classification types. Clinical diagnosis of dementia had a slightly stronger effect than Algorithmic classification on MRI variables, but none of these associations were statistically significant. The largest difference was for Cerebral Gray Matter - the average effect size for Clinical diagnosis of dementia was - 0.208, for Algorithmic classification was -0.139, so that the difference in effect sizes = 0.069 (SE of difference = 0.043, p = 0.11; note that this p value is not corrected for multiple comparisons) Comprehensive statistical significance tests for these analyses are presented in Supplemental Table 8.

**Figure 5.**
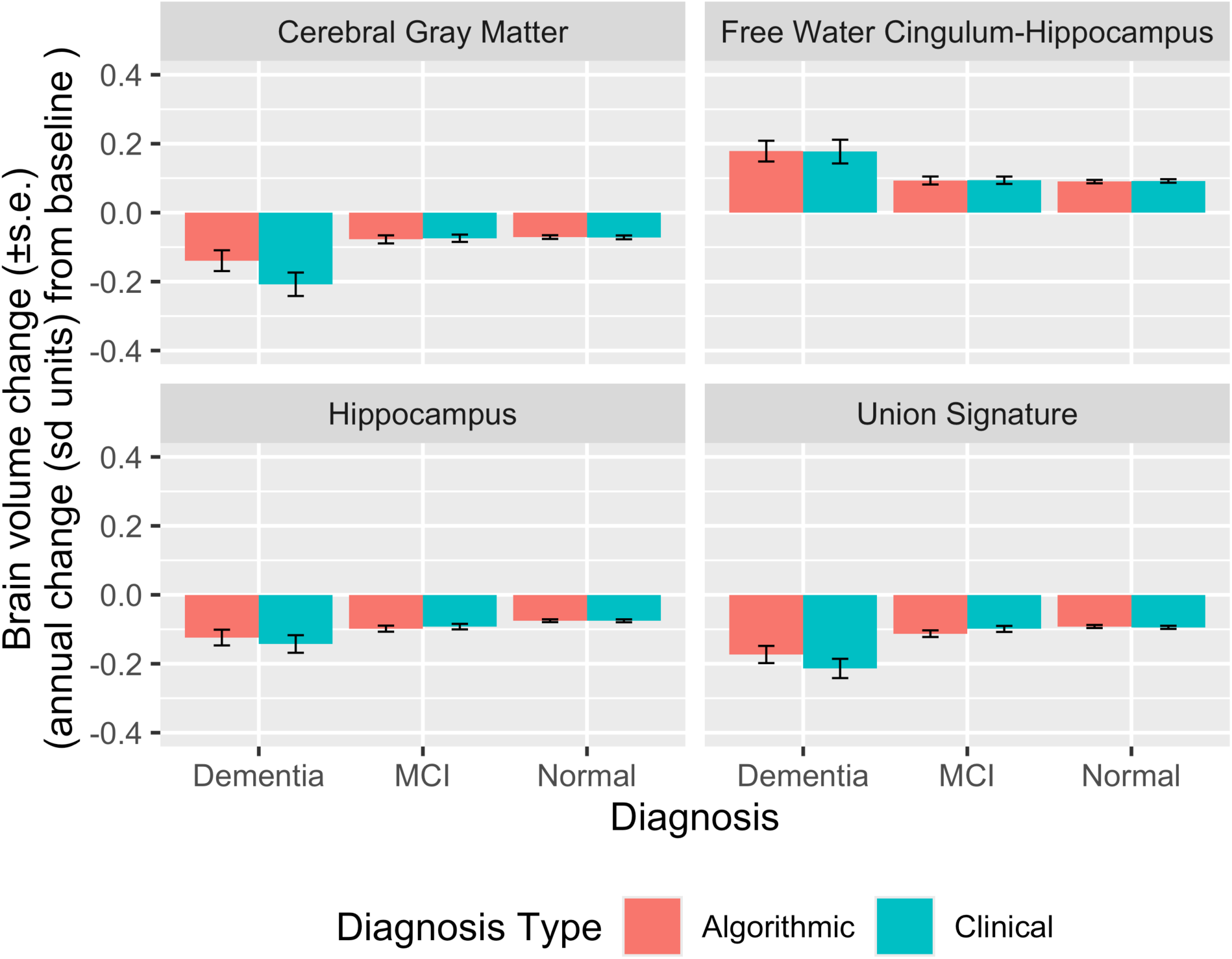
Plot of baseline diagnosis effect on brain measure change by diagnosis type - longitudinal analysis (ADRC Cohort).

In summary, results of different versions of analyses with MRI variables as longitudinal outcomes and diagnosis category-by-diagnosis/classification type interaction effects on MRI baseline and change showed a clear and consistent pattern of results. Continuous probability of a Normal diagnosis had the strongest association with longitudinal MRI. When comparing Clinical and Algorithmic categorical classification types, there were robust differences associated with diagnosis/classification category but there were negligible diagnosis/classificatin type differences within diagnosis categories.

#### 3.3.3. Longitudinal Change in Probability of Dementia in Relation to MRI Variable Baseline and Change

A final set of analyses examined how probability of dementia changed over longitudinal follow-up in relation to MRI variable baseline values and change from baseline. These analyses were restricted to the ADRC cohort. Figure 6 is a plot that shows how three diagnosis outcomes - dichotomous Clinical diagnosis of dementia (dementia vs. non-dementia), dichotomous Algorithmic classification of dementia, and continuous Algorithmic probability of dementia - are influenced by different baseline values of Union Signature and different amounts of Union Signature change. Diagnosis variables were modeled on a 0-1 probability of dementia scale. The baseline Union Signature values represented in this plot (BL) are in overall SD metrics - the average in the ADRC sample is 0. Union Signature change values (CH) are in annualized change in baseline SD Units such that 0 would indicate no change from the baseline scan and negative scores would represent decreasing union signature values (CH = - 0.25 is roughly a benchmark corresponding to -1.0 SD when union signature annualized change is transformed to a standard normal distribution, CH = -0.175 corresponds to about 0.5 SD below ADRC sample mean, and - 0.075 is approximately the mean).

**Figure 6.**
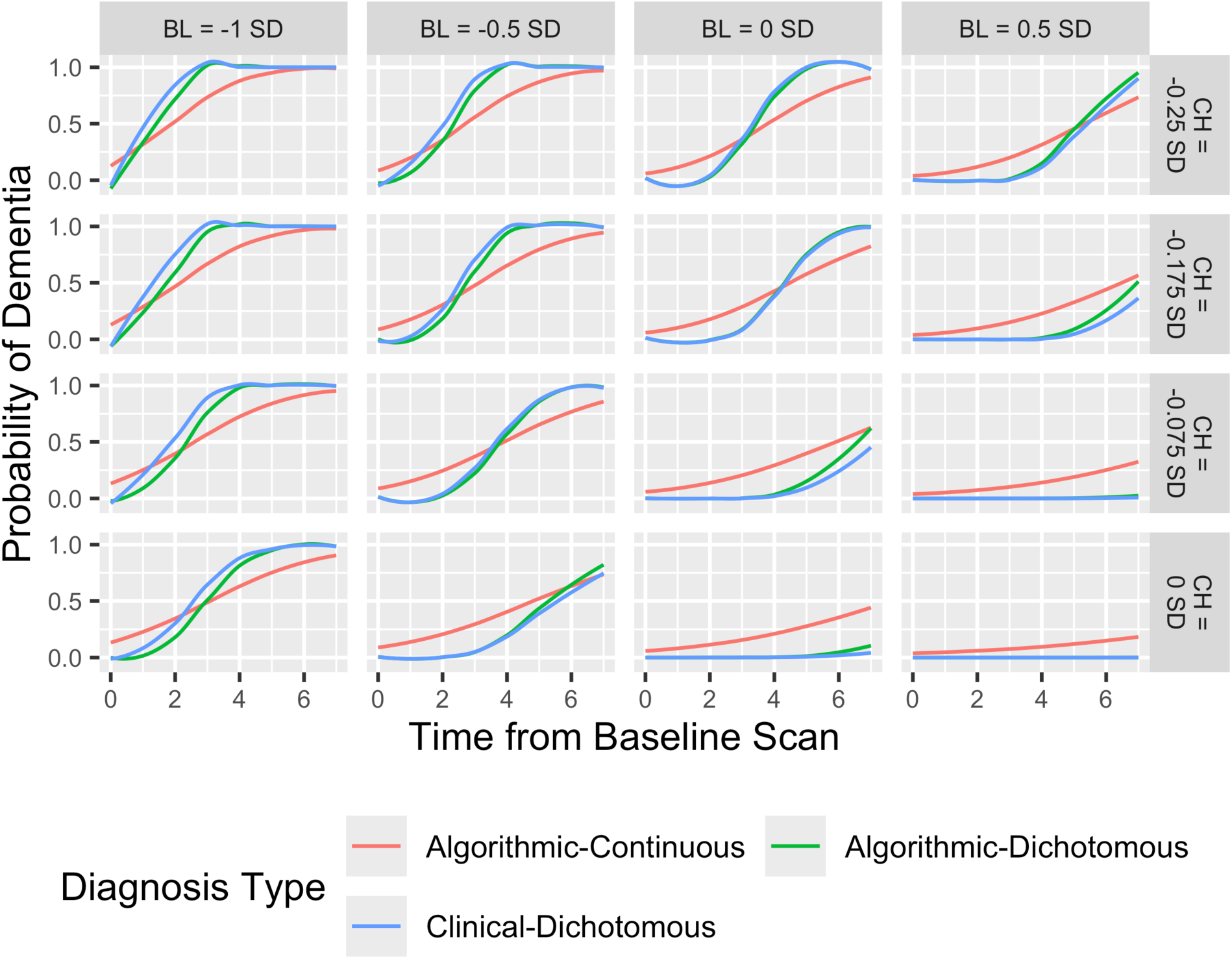
Longitudinal change in probability of dementia by union signature baseline (BL) and change (CH) and diagnosis/classification type (ADRC Cohort). [Union signature BL values are in baseline scan SD Units where 0 represents the average baseline value and the SD of baseline values = 1. Union signature CH values represent annualized change in baseline SD Units such that 0 would indicate no change from the baseline scan and negative scores would represent decreasing union signature values. CH = - 0.25 is roughly a benchmark corresponding to -1.0 SD when union signature annualized change is transformed to a standard normal distribution, CH = -0.175 corresponds to about 0.5 SD below ADRC sample mean, and -0.075 is approximately the mean.]

Several clear trends are apparent in Figure 6. 1) Probability of dementia increases for all diagnosis types as a function of lower Union Signature baseline values and greater decline in values over time. This is to be expected and provides proof of concept for this plot. 1a) Probability of dementia is low by all diagnosis types when baseline Union Signature is average or above for the sample and there is zero to mild Union Signature decline over time. 2) The probability of dementia changes more slowly over time for the continuous measure than for categorical diagnosis/classification. This too is expected given that categorical classifications artificially impose a dichotomous cutpoint on a continuous clinical process. 3) Continuous probability of dementia is more sensitive to early change (∼ 0-4 years) in Union Signature at higher BL values (0.0, 0.5). 4) Clinical diagnosis and Algorithmic classification change over time at very similar rates. There is a suggestion that clinical diagnosis changes slightly earlier than algorithmic classification at the low BL value (-1.0), but this difference stays the same across the range of CH values.

Figure 6 is useful for visualizing complex associations of change in the likelihood of a dementia diagnosis as a function of change in specific brain variables. Overall, this figure shows greater sensitivity of continuous probability of dementia (compared to Clinical diagnosis and Algorithmic classification) to early brain degeneration but a more gradual transition over time to a high probability of dementia. There were some potentially interesting differences in trajectories of Clinical diagnosis and Algorithmic classification, but these were relatively small and may not be statistically significant.

## 4. Discussion

This study examined how clinical diagnosis and algorithmic classification relate to concurrent (cross-sectional MRI measures of brain integrity) and future (progression to a more impaired diagnosis/classification and longitudinal MRI) validation criteria. Categorical clinical diagnosis was compared with two types of outcomes of a machine learning classification model, continuous probabilities of each diagnostic category/class (Normal, MCI, Dementia) and a categorical algorithmic classification that corresponded to the category with the highest probability. Continuous probability of a Normal diagnosis showed the strongest cross-sectional and longitudinal associations with MRI biomarkers while categorical clinical diagnosis, categorical algorithmic classification, and continuous probabilities of Dementia had weaker associations that did not differ among one another. When categorical clinical diagnosis was compared to categorical algorithmic classification, the overall pattern of results showed robust differences across diagnostic categories, but strikingly similar results for clinical and algorithmic classification types. Results of this study suggest that these two types of classification have very similar utility with respect to staging level of brain degeneration and predicting future clinical progression and structural brain degeneration.

A unique aspect of this study was that it compared categorical classifications with continuous probabilities of different diagnostic categories. Disease processes underlying cognitive decline in older adults, for example resulting from diseases like Alzheimer’s disease, Lewy body disease, frontotemporal degeneration, and even cerebrovascular disease, are most often gradually progressive. A continuous classification variable has a conceptual advantage in that it can capture fine grained changes in disease progression in contrast to categorical classifications which differentiate more grossly defined stages. Figure 6 illustrates these differences between continuous and categorical measures. The categorical classifications changed more slowly early and late in the disease progression process, but the transition from low to high probability was much more rapid. From a practical perspective, while grossly defined stages have well documented practical significance, they also can obscure smaller but relevant differences. An example of this is a person who is in an early period of brain degeneration. Substantial degeneration may have occurred before the threshold for a diagnosis of MCI is reached, whereas a continuous probability estimate may be able to track these changes well before a diagnosis change has occurred. Further, having information about the relative likelihood of a diagnosis can be very informative. An individual diagnosed with dementia who has a continuous probability of dementia of 0.95 is likely to be very different clinically than an individual with a diagnosis of dementia and a probability of dementia of 0.40. The former most likely has more advanced degenerative disease, and while this could be captured clinically by diagnostic sub-categories like mild, moderate, and severe dementia, this same information is provided on a more refined continuous scale by the diagnosis probability estimates. Thus, even when high quality clinical diagnosis is available, algorithmic classification probabilities may have clinical and research utility.

In this study, continuous probability of a Normal diagnosis out-performed categorical classifications and probabilities of MCI and Dementia with respect to associations with MRI measures of brain integrity. This is consistent with other research showing that continuous probabilities perform better than categorical measures derived from the continuous probabilities. [34] It isn’t entirely clear why probability of Normal would be better than probabilities of MCI or Dementia. One hypothesis to explain this is that the make-up of the study cohorts, the majority of whom had a diagnosis of Normal, biased results in favor of the Normal probability estimate. A different, methodological hypothesis is that probability of Normal captures more variance in disease progression. Supporting this hypothesis, the standard deviation for probability of Normal was larger (0.33) than for MCI (0.23) or Dementia (0.21). Further research with different samples will be needed to determine if this finding generalizes to other samples, and if so, to clarify how this relates to underlying brain degeneration.

A major finding of this study was that clinical and algorithmic classifications had very similar relationships with clinical and MRI validity criteria. However, some differences were noted. A clinical diagnosis of Normal progressed to MCI or Dementia faster than an algorithmic classification (Figure 1, left panel). This could indicate that clinical diagnosis is more sensitive to early brain degeneration. Another possible methodological explanation is that clinical diagnosis was more likely to classify borderline-impaired cases as MCI, thereby removing these cases from conversion rate comparisons because the baseline diagnosis differed across diagnosis types.

However, neither of these hypotheses are supported by associations of diagnosis with MRI variables. Future rates of brain degeneration were essentially the same in individuals with clinical and algorithmic diagnoses of Normal cognition (Figure 5) and this suggests that factors other than brain degeneration were involved.

### 4.1. Strengths and Limitations

A strength of this study was the inclusion of three cohorts of older adults that have shared cognitive assessment and MRI imaging protocols and use harmonized methods for collecting clinical information. This collective sample was unusually diverse with respect to demographic and clinical characteristics. The availability of high quality, quantitative MRI measures of brain integrity and longitudinal clinical assessment in these cohorts provided clinically and conceptually relevant validity criteria. The direct comparison of clinical and algorithmic classification with respect to concurrent and prdictive validity criteria makes an important contribution to the literature on machine learning approaches to cognitive impairment classification. There are also important limitations. Notably, the included cohorts all come from a defined region in northern California and may not be more broadly representative of California or the United States. The results of this study pertain to the specific methods used for clinical diagnosis and algorithmic classification. Of note, high quality cognitive measures, requiring 30-60 minutes of administration time, were available for algorithmic classification, and similar measures are not always available in clinical or research settings. The validation occurred in the same sample used for algorithm development, and this may have positively biased results.[35–37] However, it is not clear that this type of bias would differentially affect clinical diagnosis versus algorithmic classification. While this study makes an important contribution to understanding the utility of clinical and algorithmic classification of cognitive impairment in older adults, additional research with different samples and different methods will be important for defining both the generalizability and the limitations of the results of this study.

### 4.2. Summary and implications

This study compared clinical diagnosis and algorithmic classification of cognitive impairment with respect to ability to summarize brain integrity and predict future changes in clinical status and brain integrity. Both forms of classification had robust associations with these outcomes and this supports the clinical and research utility of both approaches and the use of algorithmic classification as a proxy for clinical diagnosis. The algorithmic classification provided a continuous measure (probability of a Normal diagnosis) that had the strongest associations with MRI based brain outcomes in this study and this represents an added value of algorithmic classification.

Clinical versus algorithmic prediction and decision making has been debated over many decades [38]. A recent review showed that clinician judgement and prognostic clinical prediction models often have similar levels of performance, but there were instances where either approach outperformed the other.[39] But this does not have to be an either-or competition. This study and the initial study from this project show how clinical and algorithmic approaches can complement one another. Two of the included cohorts (ADRC and LA90) had clinical diagnosis at every assessment but KHANDLE had clinical diagnosis for a defined subgroup of participants. Our results support use of algorithmic classification as a proxy for clinical diagnosis in KHANDLE individuals who were not clinically evaluated and also provide robust continuous classification probabilities for all assessments in all three cohorts.

Diagnosis of cognitive impairment summarizes complex patterns of information about history and progression of symptoms, clinical exam findings, and cognitive test results. Diagnosis is used to identify and stage cognitive and functional impairment and also provides information about underlying brain integrity and likelihood of future brain degeneration and clinical decline. Diagnostic criteria have been developed to promote standardization and reproducibility of diagnosis, but diagnostic decisions often require clinical judgement that is shaped by the training and experience of the diagnostician(s). Diagnostician training and experience can vary considerably across settings, as can the quality and extent of the history, exam, and test results that are available. Thus, there are many factors that can influence the quality, reproducibility, and utility of a diagnosis. Algorithmic classification can help with standardization and reproducibility but it too is dependent upon the extent and quality of input variables. Practical utility cannot be assumed for either clinical diagnosis or algorithmic classification. In this study, rather than setting clinical diagnosis as the “gold standard” and determining quality of algorithmic classification by how well it corresponded to this standard, we used independent validation criteria to examine how well both forms of classification yielded meaningful staging and prognosis information. This provides an example of a validation design that can be used to simultaneously evaluate the utility of both clinical diagnosis and algorithmic classification of cognitive impairment in a manner that improves understanding of both types of classification.

## Data Availability

All data produced in the present study are available upon reasonable request to the authors

## Notes

### Competing Interest Statement

The authors have declared no competing interest.

### Funding

- National Institute on Aging: P30AG072972, R01AG052132, R01AG056519, R01AG031563, K99AG075317

## References

[1] Mungas D, Gavett B, Rojas-Saunero LP, Zhou Y, Hayes-Larson E, Shaw C, et al. Machine learning diagnosis of cognitive impairment and dementia in harmonized older adult cohorts. Alzheimer’s & Dementia : The Journal of the Alzheimer’s Association 2025;21:e70508. 10.1002/alz.70508.

[2] Chiffi D. Clinical Reasoning: Knowledge, Uncertainty, and Values in Health Care. vol. 58. Cham: Springer International Publishing; 2021. 10.1007/978-3-030-59094-9.

[3] Newman TB, Kohn MA. Evidence-Based Diagnosis: An Introduction to Clinical Epidemiology. Cambridge, UK: dge University Press; 2020.

[4] Atri A, Dickerson BC, Clevenger C, Karlawish J, Knopman D, Lin P-J, et al. Alzheimer’s Association clinical practice guideline for the Diagnostic Evaluation, Testing, Counseling, and Disclosure of Suspected Alzheimer’s Disease and Related Disorders (DETeCD-ADRD): Executive summary of recommendations for primary care. Alzheimer’s & Dementia 2025;21:e14333. 10.1002/alz.14333.

[5] Livingston G, Huntley J, Liu KY, Costafreda SG, Selbæk G, Alladi S, et al. Dementia prevention, intervention, and care: 2024 report of the Lancet standing Commission. The Lancet 2024;404:572–628. 10.1016/S0140-6736(24)01296-0.

[6] Vemuri P, Wiste HJ, Weigand SD, Shaw LM, Trojanowski JQ, Weiner MW, et al. MRI and CSF biomarkers in normal, MCI, and AD subjects: Predicting future clinical change. Neurology 2009;73:294–301. 10.1212/WNL.0b013e3181af79fb.

[7] Fletcher E, Knaack A, Singh B, Lloyd E, Wu E, Carmichael O, et al. Combining Boundary-Based Methods With Tensor-Based Morphometry in the Measurement of Longitudinal Brain Change. IEEE Transactions on Medical Imaging 2013;32:223–36. 10.1109/TMI.2012.2220153.

[8] Chen Y, Denny KG, Harvey D, Farias ST, Mungas D, DeCarli C, et al. Progression from normal cognition to mild cognitive impairment in a diverse clinic-based and community-based elderly cohort. Alzheimer’s & Dementia 2017;13:399–405. 10.1016/j.jalz.2016.07.151.

[9] Farias ST, Mungas D, Reed BR, Harvey D, DeCarli C. Progression of mild cognitive impairment to dementia in clinic-vs community-based cohorts. Arch Neurol 2009;66:1151–7. 10.1001/archneurol.2009.106.

[10] Mungas D, Beckett L, Harvey D, Tomaszewski Farias S, Reed B, Carmichael O, et al. Heterogeneity of cognitive trajectories in diverse older persons. Psychology and Aging 2010;25. 10.1037/a0019502.

[11] Martin SA, Townend FJ, Barkhof F, Cole JH. Interpretable machine learning for dementia: A systematic review. Alzheimer’s & Dementia 2023;19:2135–49. 10.1002/alz.12948.

[12] Hinton L, Carter K, Reed BR, Beckett L, Lara E, DeCarli C, et al. Recruitment of a community-based cohort for research on diversity and risk of dementia. Alzheimer Dis Assoc Disord 2010;24:234–41. 10.1097/WAD.0b013e3181c1ee01.

[13] Besser L, Kukull W, Knopman DS, Chui H, Galasko D, Weintraub S, et al. Version 3 of the National Alzheimer’s Coordinating Center’s Uniform Data Set. Alzheimer Disease and Associated Disorders 2018;32:351–8. 10.1097/WAD.0000000000000279.

[14] Weintraub S, Besser L, Dodge HH, Teylan M, Ferris S, Goldstein FC, et al. Version 3 of the Alzheimer Disease Centers’ Neuropsychological Test Battery in the Uniform Data Set (UDS). Alzheimer Disease and Associated Disorders 2018;32:10–7. 10.1097/WAD.0000000000000223.

[15] Teng EL, Chui HC. The Modified Mini-Mental State (3MS) Examination. Journal of Clinical Psychiatry 1987;48:314–8.

[16] Morris JC, Weintraub S, Chui HC, Cummings J, Decarli C, Ferris S, et al. The Uniform Data Set (UDS): Clinical and cognitive variables and descriptive data from Alzheimer Disease Centers. Alzheimer’s Disease and Associated Disorders 2006;20:210–6. https://doi.org/10.1097/01.wad.0000213865.09806.9200002093-200610000-00007 [pii].

[17] Morris JC. The Clinical Dementia Rating (CDR): Current version and scoring rules. Neurology 1993;43:2412–4.

[18] Mungas D, Reed BR, Crane PK, Haan MN, Gonzalez H. Spanish and English Neuropsychological Assessment Scales (SENAS): Further development and psychometric characteristics. Psychol Assess 2004;16:347–59. 10.1037/1040-3590.16.4.347.

[19] Hernandez Saucedo H, Whitmer RA, Glymour M, DeCarli C, Mayeda E-R, Gilsanz P, et al. Measuring Cognitive Health in Ethnically Diverse Older Adults. The Journals of Gerontology Series B, Psychological Sciences and Social Sciences 2022;77:261–71. 10.1093/geronb/gbab062.

[20] Farias ST, Mungas D, Reed BR, Cahn-Weiner D, Jagust W, Baynes K, et al. The measurement of everyday cognition (ECog): Scale development and psychometric properties. Neuropsychology 2008;22:531–44. 10.1037/0894-4105.22.4.531.

[21] Fletcher E, DeCarli C, Fan AP, Knaack A. Convolutional Neural Net Learning Can Achieve Production-Level Brain Segmentation in Structural Magnetic Resonance Imaging. Frontiers in Neuroscience 2021;15:683426. 10.3389/fnins.2021.683426.

[22] Fletcher E, Carmichael O, Pasternak O, Maier-Hein KH, DeCarli C. Early Brain Loss in Circuits Affected by Alzheimer’s Disease is Predicted by Fornix Microstructure but may be Independent of Gray Matter. Front Aging Neurosci 2014;6:1–9. 10.3389/fnagi.2014.00106.

[23] Maillard P, Fletcher E, Singh B, Martinez O, Johnson DK, Olichney JM, et al. Cerebral white matter free water: A sensitive biomarker of cognition and function. Neurology 2019;92:e2221–31. 10.1212/WNL.0000000000007449.

[24] Fletcher E, Gavett B, Harvey D, Farias ST, Olichney J, Beckett L, et al. Brain volume change and cognitive trajectories in aging. Neuropsychology 2018;32:436–49. 10.1037/neu0000447.

[25] Lee DY, Fletcher E, Martinez O, Zozulya N, Kim J, Tran J, et al. Vascular and degenerative processes differentially affect regional interhemispheric connections in normal aging, mild cognitive impairment, and Alzheimer disease. Stroke 2010;41:1791–7. 10.1161/STROKEAHA.110.582163.

[26] Fletcher E, Gavett B, Crane P, Soldan A, Hohman T, Farias S, et al. A robust brain signature region approach for episodic memory performance in older adults. Brain : A Journal of Neurology 2021;144:1089–102. 10.1093/brain/awab007.

[27] Fletcher E, Farias S, DeCarli C, Gavett B, Widaman K, De Leon F, et al. Toward a statistical validation of brain signatures as robust measures of behavioral substrates. Human Brain Mapping 2023;44:3094–111. 10.1002/hbm.26265.

[28] Fletcher E, Gavett B, Farias ST, Widaman K, Whitmer R, Fan AP, et al. A data-driven, multi-domain brain gray matter signature as a powerful biomarker associated with several clinical outcomes. Alzheimer’s & Dementia (Amsterdam, Netherlands) n.d.;16:e70026. 10.1002/dad2.70026.

[29] Pasternak O, Sochen N, Gur Y, Intrator N, Assaf Y. Free water elimination and mapping from diffusion MRI. Magnetic Resonance in Medicine 2009;62:717–30. 10.1002/mrm.22055.

[30] Johnson WE, Li C, Rabinovic A. Adjusting batch effects in microarray expression data using empirical Bayes methods. Biostatistics 2007;8:118–27. 10.1093/biostatistics/kxj037.

[31] Tassi E, Bianchi AM, Calesella F, Vai B, Bellani M, Nenadić I, et al. Assessment of ComBat Harmonization Performance on Structural Magnetic Resonance Imaging Measurements. Human Brain Mapping 2024;45:e70085. 10.1002/hbm.70085.

[32] Beer JC, Tustison NJ, Cook PA, Davatzikos C, Sheline YI, Shinohara RT, et al. Longitudinal ComBat: A method for harmonizing longitudinal multi-scanner imaging data. NeuroImage 2020;220:117129. 10.1016/j.neuroimage.2020.117129.

[33] Blom G. Statistical estimates and transformed beta-variables. New York: Wiley; 1958.

[34] Blythe R, Parsons R, Ong ME, Barnett A. Continuous predicted risks should be retained when deploying clinical prediction models. Journal of Clinical Epidemiology 2025;188. 10.1016/j.jclinepi.2025.112009.

[35] de Hond AAH, Shah VB, Kant IMJ, Van Calster B, Steyerberg EW, Hernandez-Boussard T. Perspectives on validation of clinical predictive algorithms. Npj Digital Medicine 2023;6:86. 10.1038/s41746-023-00832-9.

[36] Roi-Teeuw HM la, Royen FS van, Hond A de, Zahra A, Vries S de, Bartels R, et al. Don’t be misled: 3 misconceptions about external validation of clinical prediction models. Journal of Clinical Epidemiology 2024;172. 10.1016/j.jclinepi.2024.111387.

[37] Kaul T, Kellerhuis BE, Damen JAA, Schuit E, Jenniskens K, Smeden M van, et al. Methodological quality assessment tools for diagnosis and prognosis research: Overview and guidance. Journal of Clinical Epidemiology 2025;177. 10.1016/j.jclinepi.2024.111609.

[38] Meehl PE. Clinical versus statistical prediction: A theoretical analysis and a review of the evidence. Minneapolis, MN, US: University of Minnesota Press; 1954. 10.1037/11281-000.

[39] Colunga-Lozano LE, Foroutan F, Rayner D, Luca CD, Hernández-Wolters B, Couban R, et al. Clinical judgment shows similar and sometimes superior discrimination compared to prognostic clinical prediction models: A systematic review. Journal of Clinical Epidemiology 2024;165. 10.1016/j.jclinepi.2023.10.016.

