## Appendix and Supplemental Tables for "Validation of clinical diagnosis and machine learning classification of cognitive impairment"

### A.1. Methods

#### A.1.1. MRI Adjustment for Field Strength and DTI Directions

MRI scans for ADRC cohort were on a 1.5 Tesla (1.5T) scanner through 2016 but were on a 3T scanner beginning in 2017. The number of directions involved in diffusion tensor imaging (DTI) scans changed with the scanner change initiated in 2017. We adjusted union signature, total gray matter volume, and hippocampal volume for scanner field strength and adjusted free water cingulum-hippocampus for scanner field strength and and number directions in the DTI scan.

The relevance of adjustment for scanner properties was evaluated in the ADRC cohort. In this cohort, 58.8% of those whose first scan was a 1.5T scan were cognitively Normal (7.2% had Dementia) compared to 88.7% of those who received a 3T first scan (0% had Dementia). Robust cross-sectional field strength effects were present for some MRI measures. For union signature for example, field strength explained 13.9% of the variance in the baseline, raw union signature measurement. Differences in measurement due to clinical/brain health are of direct interest for this study, but these effects are potentially confounded with methodological effects due to different levels of resolution of the MRI images associated with field strength and DTI directions.

We harmonized scans using the ComBat ("combating batch effects when combining batches") method, which was originally developed for genomic data [1]. This method has been used to harmonize both structural MRI and DTI [2]. We used a recent adaption of COMBAT for longitudinal data developed by Beer [3] that is implemented in the R longCombat package (version 0.0.0.90000). The general longCombat approach uses a mixed effects model to harmonize scan scores across batches. For field strength adjustment, batches corresponded to scanner field strength. MRI variables (features) were regressed on biological variables known to affect scan results (typically age and sex), time from first scan, and covariates. We chose baseline clinical diagnosis (not time-varying) and continuous probability of Normal (time varying) as covariates of interest. An intercept random effect was specified to account for within-person correlation of MRI features across repeated scans.

Total gray matter and hippocampal volumes were first residualized for intracranial volume. These residualized volumes, union signature, and freewater cingulum-hippocampus were then Blom standardized [4]. longCombat requires balanced data (no missing values) so we used the R mice package (version 3.16.0) to perform multiple imputation on a dataset that included the 4 standardized MRI features along with age, sex, baseline clinical diagnosis and continuous probability of a Normal diagnosis. Missing values were replaced with their respective medians across 25 imputations. For the freewater DTI variable, batch was defined by the combination of scanner field strength and the number of DTI direction for the scan. For the other 3 variables, batch was defined by field strength. The formula for the longCombat model was:

$$time\_sc + age + female + Dx_{bl} + prNorm + time\_sc * female + time\_sc * Dx_{bl}$$

where time_sc = time in years from the first scan, age = age in years (time-varying across longitudinal scans), female = female sex (1 if female, 0 otherwise, time-invariant), $Dx_{bl}$ = clinical diagnosis at time of first scan (time-invariant), and $prNorm$ = continuous probability of a Normal diagnosis (time-varying). The interactions of time_sc with female and $Dx_{bl}$ were also included. ranef was specified as (1|id) - intercept random effect. longCombat generated values of MRI features harmonized across batches and these variables were then Blom standardized to create a final set of analysis variables that were normally distributed and had a mean of 0 and SD of 1 in the overall longitudinal MRI sample.

The effectiveness of the longCombat harmonization was evaluated using a mixed effects model symbolically represented by:

$$MRI_{variable} \sim \beta_0 + \beta_T * time\_sc + \beta_D * Dx_{bl} + \beta_N * prNorm + \beta_{FS} * FS + \beta_{Dir} * DTI_{dirs} + \beta_{TxD} * time\_sc * Dx_{bl} + (1|id) + \epsilon$$

where $MRI_{variable}$ = the MRI variable, time_sc = time from 1st scan, $Dx_{bl}$ = clinical diagnosis at time of first scan, $prNorm$ = probability of a Normal diagnosis, FS = scanner field strength (0 if 1.5T, 1 if 3T), $DTI_{dirs}$ = number of DTI directions, $\beta_0$, $\beta_T$,$\beta_D$,$\beta_N$,$\beta_{FS}$, $\beta_{Dir}$, and $\beta_{TxD}$ are regression coefficients, (1|id) specifies an intercept random effect, and $\epsilon$ = random error. This model was estimated for unadjusted MRI values and for MRI values that were adjusted using longCombat.

Supplemental Table 1 presents coefficients from the mixed effects regression models evaluating field strength and DTI direction effects in the longitudinal models that accounted for baseline clinical diagnosis and time-varying probability of Normal diagnosis effects on MRI variables. The Field Strength and DTI Directions (for freewater cingulum-hippocampus) effects show how strongly these MRI methodological parameters impacted MRI measures independent of diagnosis variables. Field Strength and DTI Direction effects were small and non-significant for the longCombat adjusted scores, but were larger and statistically significant for undjusted scores for all MRI measure except hippocampus. Associations of the clinical and algorithmic diagnosis/classification variables were very similar for adjusted and unadjusted MRI variables.

### References


[1] Johnson WE, Li C, Rabinovic A. Adjusting batch effects in microarray expression data using empirical Bayes methods. Biostatistics 2007;8:118–27. https://doi.org/10.1093/biostatistics/kxj037.

[2] Tassi E, Bianchi AM, Calesella F, Vai B, Bellani M, Nenadić I, et al. Assessment of ComBat Harmonization Performance on Structural Magnetic Resonance Imaging Measurements. Human Brain Mapping 2024;45:e70085. https://doi.org/10.1002/hbm.70085.

[3] Beer JC, Tustison NJ, Cook PA, Davatzikos C, Sheline YI, Shinohara RT, et al. Longitudinal ComBat: A method for harmonizing longitudinal multi-scanner imaging data. NeuroImage 2020;220:117129. https://doi.org/10.1016/j.neuroimage.2020.117129.

[4] Blom G. Statistical estimates and transformed beta-variables. New York: Wiley; 1958.

Supplemental Table 1. Effects of field strength, DTI directions, and diagnoses on batch adjusted and unadjusted MRI variables. [Key - effect: Intercept = average value at time of baseline scan for “Normal” baseline clinical diagnosis, Time from 1st Scan = time (years) from 1st scan, Dx_bl MCI = average value at time of baseline scan for “MCI” baseline clinical diagnosis, Dx_bl Dementia = average value at time of baseline scan for “Dementia” baseline clinical diagnosis, Probability Normal Dx = algorithmically predicted probability of a Normal clinical diagnosis (time-varying), Field Strength = scanner field strength (time-varying), DTI Directions = number of DTI directions (time-varying), Time x Dx_bl MCI = interaction of Time from 1st Scan and Dx_bl MCI, Time x Dx_bl MCI = interaction of Time from 1st Scan and Dx_bl Dementia]

| mri_variable | adjustment | effect | estimate | SE | t | p |
|---|---|---|---|---|---|---|
| Freewater Cingulum Hippocampus | Combat Adjusted | Intercept | 0.38 | 0.14 | 2.77 | 0.01 |
| Freewater Cingulum Hippocampus | Unadjusted | Intercept | 1.52 | 0.10 | 15.85 | 0.00 |
| Freewater Cingulum Hippocampus | Combat Adjusted | Time from 1st Scan | 0.06 | 0.01 | 8.57 | 0.00 |
| Freewater Cingulum Hippocampus | Unadjusted | Time from 1st Scan | 0.05 | 0.00 | 9.53 | 0.00 |
| Freewater Cingulum Hippocampus | Combat Adjusted | Dx_bl MCI | 0.11 | 0.10 | 1.03 | 0.30 |
| Freewater Cingulum Hippocampus | Unadjusted | Dx_bl MCI | 0.13 | 0.07 | 1.75 | 0.08 |
| Freewater Cingulum Hippocampus | Combat Adjusted | Dx_bl Dementia | 0.07 | 0.19 | 0.35 | 0.73 |
| Freewater Cingulum Hippocampus | Unadjusted | Dx_bl Dementia | 0.12 | 0.14 | 0.84 | 0.40 |
| Freewater Cingulum Hippocampus | Combat Adjusted | Probability Normal Dx | -0.93 | 0.09 | -10.29 | 0.00 |
| Freewater Cingulum Hippocampus | Unadjusted | Probability Normal Dx | -0.67 | 0.06 | -10.48 | 0.00 |

| | | | | | | |
|---|---|---|---|---|---|---|
| Freewater Cingulum Hippocampus | Combat Adjusted | Field Strength | 0.09 | 0.14 | 0.69 | 0.49 |
| Freewater Cingulum Hippocampus | Unadjusted | Field Strength | -0.51 | 0.10 | -5.33 | 0.00 |
| Freewater Cingulum Hippocampus | Combat Adjusted | DTI Directions | 0.00 | 0.00 | -0.31 | 0.75 |
| Freewater Cingulum Hippocampus | Unadjusted | DTI Directions | -0.02 | 0.00 | -9.66 | 0.00 |
| Freewater Cingulum Hippocampus | Combat Adjusted | Time x Dx_bl MCI | 0.01 | 0.01 | 0.92 | 0.36 |
| Freewater Cingulum Hippocampus | Unadjusted | Time x Dx_bl MCI | 0.01 | 0.01 | 1.10 | 0.27 |
| Freewater Cingulum Hippocampus | Combat Adjusted | Time x Dx_bl Dementia | 0.10 | 0.04 | 2.41 | 0.02 |
| Freewater Cingulum Hippocampus | Unadjusted | Time x Dx_bl Dementia | 0.08 | 0.03 | 2.71 | 0.01 |
| Hippocampus | Combat Adjusted | Intercept | -0.12 | 0.08 | -1.50 | 0.13 |
| Hippocampus | Unadjusted | Intercept | -0.01 | 0.07 | -0.10 | 0.92 |
| Hippocampus | Combat Adjusted | Time from 1st Scan | -0.06 | 0.01 | -10.55 | 0.00 |
| Hippocampus | Unadjusted | Time from 1st Scan | -0.06 | 0.00 | -15.38 | 0.00 |
| Hippocampus | Combat Adjusted | Dx_bl MCI | -0.43 | 0.10 | -4.41 | 0.00 |
| Hippocampus | Unadjusted | Dx_bl MCI | -0.45 | 0.09 | -5.19 | 0.00 |
| Hippocampus | Combat Adjusted | Dx_bl Dementia | -0.95 | 0.19 | -5.04 | 0.00 |
| Hippocampus | Unadjusted | Dx_bl Dementia | -1.06 | 0.18 | -5.79 | 0.00 |

| | | | | | | |
|---|---|---|---|---|---|---|
| Hippocampus | Combat Adjusted | Probability Normal Dx | 0.66 | 0.08 | 8.13 | 0.00 |
| Hippocampus | Unadjusted | Probability Normal Dx | 0.54 | 0.06 | 9.56 | 0.00 |
| Hippocampus | Combat Adjusted | Field Strength | 0.08 | 0.05 | 1.55 | 0.12 |
| Hippocampus | Unadjusted | Field Strength | 0.07 | 0.04 | 1.79 | 0.07 |
| Hippocampus | Combat Adjusted | Time x Dx_bl MCI | -0.02 | 0.01 | -2.24 | 0.03 |
| Hippocampus | Unadjusted | Time x Dx_bl MCI | -0.03 | 0.01 | -3.45 | 0.00 |
| Hippocampus | Combat Adjusted | Time x Dx_bl Dementia | -0.07 | 0.03 | -2.18 | 0.03 |
| Hippocampus | Unadjusted | Time x Dx_bl Dementia | -0.08 | 0.02 | -4.09 | 0.00 |
| Total Gray Matter | Combat Adjusted | Intercept | -0.20 | 0.09 | -2.14 | 0.03 |
| Total Gray Matter | Unadjusted | Intercept | -0.40 | 0.09 | -4.70 | 0.00 |
| Total Gray Matter | Combat Adjusted | Time from 1st Scan | -0.05 | 0.01 | -6.93 | 0.00 |
| Total Gray Matter | Unadjusted | Time from 1st Scan | -0.05 | 0.01 | -7.64 | 0.00 |
| Total Gray Matter | Combat Adjusted | Dx_bl MCI | -0.30 | 0.10 | -2.90 | 0.00 |
| Total Gray Matter | Unadjusted | Dx_bl MCI | -0.29 | 0.10 | -2.99 | 0.00 |
| Total Gray Matter | Combat Adjusted | Dx_bl Dementia | -0.19 | 0.19 | -0.97 | 0.33 |
| Total Gray Matter | Unadjusted | Dx_bl Dementia | -0.23 | 0.18 | -1.28 | 0.20 |

| | | | | | | |
|---|---|---|---|---|---|---|
| Total Gray Matter | Combat Adjusted | Probability Normal Dx | 0.76 | 0.10 | 7.81 | 0.00 |
| Total Gray Matter | Unadjusted | Probability Normal Dx | 0.71 | 0.09 | 8.08 | 0.00 |
| Total Gray Matter | Combat Adjusted | Field Strength | -0.02 | 0.06 | -0.31 | 0.76 |
| Total Gray Matter | Unadjusted | Field Strength | 0.47 | 0.06 | 8.39 | 0.00 |
| Total Gray Matter | Combat Adjusted | Time x Dx_bl MCI | -0.01 | 0.01 | -0.77 | 0.44 |
| Total Gray Matter | Unadjusted | Time x Dx_bl MCI | -0.02 | 0.01 | -1.31 | 0.19 |
| Total Gray Matter | Combat Adjusted | Time x Dx_bl Dementia | -0.14 | 0.04 | -3.34 | 0.00 |
| Total Gray Matter | Unadjusted | Time x Dx_bl Dementia | -0.13 | 0.03 | -3.74 | 0.00 |
| Union Signature | Combat Adjusted | Intercept | -0.23 | 0.09 | -2.71 | 0.01 |
| Union Signature | Unadjusted | Intercept | -0.41 | 0.07 | -5.61 | 0.00 |
| Union Signature | Combat Adjusted | Time from 1st Scan | -0.07 | 0.01 | -11.00 | 0.00 |
| Union Signature | Unadjusted | Time from 1st Scan | -0.07 | 0.01 | -14.18 | 0.00 |
| Union Signature | Combat Adjusted | Dx_bl MCI | -0.34 | 0.10 | -3.62 | 0.00 |
| Union Signature | Unadjusted | Dx_bl MCI | -0.38 | 0.09 | -4.40 | 0.00 |
| Union Signature | Combat Adjusted | Dx_bl Dementia | -0.57 | 0.18 | -3.14 | 0.00 |
| Union Signature | Unadjusted | Dx_bl Dementia | -0.67 | 0.17 | -4.01 | 0.00 |

| | | | | | | |
|---|---|---|---|---|---|---|
| Union Signature | Combat Adjusted | Probability Normal Dx | 0.85 | 0.09 | 9.78 | 0.00 |
| Union Signature | Unadjusted | Probability Normal Dx | 0.66 | 0.07 | 9.29 | 0.00 |
| Union Signature | Combat Adjusted | Field Strength | 0.04 | 0.05 | 0.81 | 0.42 |
| Union Signature | Unadjusted | Field Strength | 0.68 | 0.05 | 14.66 | 0.00 |
| Union Signature | Combat Adjusted | Time x Dx_bl MCI | -0.01 | 0.01 | -1.04 | 0.30 |
| Union Signature | Unadjusted | Time x Dx_bl MCI | -0.02 | 0.01 | -1.88 | 0.06 |
| Union Signature | Combat Adjusted | Time x Dx_bl Dementia | -0.13 | 0.03 | -3.67 | 0.00 |
| Union Signature | Unadjusted | Time x Dx_bl Dementia | -0.11 | 0.03 | -4.22 | 0.00 |

Supplemental Table 2. Sample characteristics by analysis type. Column totals are numbers of individuals contributing to each analysis.

| | Diagnosis Conversion - MCI at Baseline (N=249) | Diagnosis Conversion - Normal at Baseline (N=1009) | MRI - Cross-sectional (N=1580) | MRI - Longitudinal (N=528) |
|---|---|---|---|---|
| **sex/gender** | | | | |
| Mean (SD) | 1.55 (±0.498) | 1.64 (±0.481) | 1.59 (±0.492) | 1.61 (±0.488) |
| **age (baseline) (years)** | | | | |
| Mean (SD) | 82.3 (±8.79) | 80.0 (±10.2) | 77.2 (±8.70) | 74.1 (±7.06) |
| **education (years)** | | | | |
| Mean (SD) | 14.0 (±3.58) | 14.6 (±3.37) | 13.8 (±4.20) | 13.9 (±4.29) |
| **race/ethnicity** | | | | |
| Asian | 29 (11.9%) | 177 (17.7%) | 160 (10.2%) | 23 (4.4%) |
| Black | 68 (27.9%) | 226 (22.6%) | 377 (24.1%) | 112 (21.3%) |
| LatinX | 35 (14.3%) | 237 (23.7%) | 383 (24.5%) | 135 (25.6%) |
| White | 112 (45.9%) | 355 (35.5%) | 638 (40.8%) | 256 (48.6%) |
| Native American | 0 (0%) | 4 (0.4%) | 1 (0.1%) | 0 (0%) |
| NativeAmer | 0 (0%) | 1 (0.1%) | 4 (0.3%) | 1 (0.2%) |
| **diagnosis (baseline)** | | | | |
| Normal | 0 (0%) | 1009 (100%) | 1012 (64.1%) | 351 (66.5%) |
| MCI | 249 (100%) | 0 (0%) | 427 (27.0%) | 147 (27.8%) |
| Dementia | 0 (0%) | 0 (0%) | 141 (8.9%) | 30 (5.7%) |
| **study** | | | | |
| ADC | 120 (48.2%) | 388 (38.5%) | 1095 (69.3%) | 528 (100%) |
| KHANDLE | 61 (24.5%) | 266 (26.4%) | 282 (17.8%) | 0 (0%) |
| LA90 | 68 (27.3%) | 355 (35.2%) | 203 (12.8%) | 0 (0%) |
| **N assessments** | | | | |
| Mean (SD) | 4.35 (±2.07) | 5.36 (±2.81) | 4.19 (±3.11) | 6.62 (±3.55) |

| | Diagnosis Conversion - MCI at Baseline (N=249) | Diagnosis Conversion - Normal at Baseline (N=1009) | MRI - Cross-sectional (N=1580) | MRI - Longitudinal (N=528) |
|---|---|---|---|---|
| **follow-up time (years)** | | | | |
| Mean (SD) | 4.04 (±2.31) | 5.18 (±3.63) | 4.18 (±3.88) | 7.30 (±4.47) |

Supplemental Table 3. Model performance comparison for MRI brain measure associations with diagnosis variables, cross-sectional. [Key - Diagnosis/Classification Variable: Clinical Diagnosis = Clinical categorical diagnosis, Algorithmic Classification = Algorithmic categorical classification, prob(Normal) = Algorithmically predicted probability of a Normal clinical diagnosis, prob(MCI) = Algorithmically predicted probability of a MCI clinical diagnosis, prob(Dementia) = Algorithmically predicted probability of a Dementia clinical diagnosis; AIC = Akaike Information Criterion, BIC = Bayesian Information Criterion, R2 = R-Squared, RMSE = Root Mean Square Error]

| MRI Measure | Diagnosis/Classification Variable | AIC | BIC | R2 | RMSE |
|---|---|---|---|---|---|
| Union Signature | Clinical Diagnosis | 4473 | 4495 | 0.116 | 0.947 |
| Union Signature | Algorithmic Classification | 4480 | 4502 | 0.112 | 0.949 |
| Union Signature | prob(Normal) | 4393 | 4409 | 0.157 | 0.925 |
| Union Signature | prob(MCI) | 4526 | 4543 | 0.086 | 0.963 |
| Union Signature | prob(Dementia) | 4505 | 4521 | 0.098 | 0.957 |
| Freewater Cingulum-Hippocampus | Clinical Diagnosis | 3395 | 3416 | 0.084 | 0.932 |
| Freewater Cingulum-Hippocampus | Algorithmic Classification | 3392 | 3413 | 0.087 | 0.931 |
| Freewater Cingulum-Hippocampus | prob(Normal) | 3334 | 3350 | 0.126 | 0.91 |
| Freewater Cingulum-Hippocampus | prob(MCI) | 3412 | 3427 | 0.071 | 0.939 |
| Freewater Cingulum-Hippocampus | prob(Dementia) | 3401 | 3417 | 0.078 | 0.935 |
| Hippocampus | Clinical Diagnosis | 4384 | 4405 | 0.138 | 0.922 |
| Hippocampus | Algorithmic Classification | 4395 | 4417 | 0.132 | 0.925 |
| Hippocampus | prob(Normal) | 4318 | 4335 | 0.17 | 0.904 |
| Hippocampus | prob(MCI) | 4446 | 4463 | 0.103 | 0.94 |
| Hippocampus | prob(Dementia) | 4466 | 4482 | 0.092 | 0.946 |

| | | | | | |
|---|---|---|---|---|---|
| Cerebral Gray Matter | Clinical Diagnosis | 4663 | 4685 | 0.063 | 1.004 |
| Cerebral Gray Matter | Algorithmic Classification | 4658 | 4680 | 0.066 | 1.002 |
| Cerebral Gray Matter | prob(Normal) | 4626 | 4642 | 0.083 | 0.993 |
| Cerebral Gray Matter | prob(MCI) | 4702 | 4718 | 0.039 | 1.016 |
| Cerebral Gray Matter | prob(Dementia) | 4665 | 4682 | 0.06 | 1.005 |

Supplemental Table 4. Model performance comparison for longitudinal imaging measure associations with diagnosis variables (time-varying, ADRC Cohort). [Key - Diagnosis/Classification Variable: Clinical Diagnosis = Clinical categorical diagnosis, Algorithmic Classification = Algorithmic categorical classification, prob(Normal) = Algorithmically predicted probability of a Normal clinical diagnosis, prob(MCI) = Algorithmically predicted probability of a MCI clinical diagnosis, prob(Dementia) = Algorithmically predicted probability of a Dementia clinical diagnosis; AIC = Akaike Information Criterion, BIC = Bayesian Information Criterion, R2_marginal = Marginal R-Squared]

| MRI Variable | Diagnosis/Classification Variable | AIC | BIC | R2_marginal |
|---|---|---|---|---|
| Union Signature | Clinical Diagnosis | 3098 | 3130 | 0.176 |
| Union Signature | Algorithmic Classification | 3153 | 3184 | 0.196 |
| Union Signature | prob(Normal) | 3134 | 3160 | 0.243 |
| Union Signature | prob(MCI) | 3339 | 3365 | 0.084 |
| Union Signature | prob(Dementia) | 3168 | 3194 | 0.162 |
| Freewater Cingulum-Hippocampus | Clinical Diagnosis | 2549 | 2580 | 0.186 |
| Freewater Cingulum-Hippocampus | Algorithmic Classification | 2587 | 2618 | 0.18 |
| Freewater Cingulum-Hippocampus | prob(Normal) | 2554 | 2579 | 0.226 |
| Freewater Cingulum-Hippocampus | prob(MCI) | 2725 | 2750 | 0.09 |
| Freewater Cingulum-Hippocampus | prob(Dementia) | 2571 | 2597 | 0.17 |
| Hippocampus | Clinical Diagnosis | 2997 | 3029 | 0.127 |
| Hippocampus | Algorithmic Classification | 3028 | 3059 | 0.126 |
| Hippocampus | prob(Normal) | 3006 | 3032 | 0.164 |
| Hippocampus | prob(MCI) | 3161 | 3187 | 0.059 |

| | | | | |
|---|---|---|---|---|
| Hippocampus | prob(Dementia) | 3040 | 3066 | 0.1 |
| Cerebral Gray Matter | Clinical Diagnosis | 3397 | 3428 | 0.134 |
| Cerebral Gray Matter | Algorithmic Classification | 3416 | 3447 | 0.155 |
| Cerebral Gray Matter | prob(Normal) | 3422 | 3448 | 0.17 |
| Cerebral Gray Matter | prob(MCI) | 3569 | 3596 | 0.052 |
| Cerebral Gray Matter | prob(Dementia) | 3428 | 3454 | 0.131 |

Supplemental Table 5. Time-varying diagnosis/classification effect estimates by diagnosis type on brain measures in longitudinal models (ADRC cohort). (Diagnosis/classification is time-varying independent variable in mixed effects longitudinal models of longitudinal change in brain measures. Diagnosis/classification effects combine between-person diagnosis group differences in brain measures and within-person change associated with change in diagnosis/classification. p values are not corrected for multiple comparisons.) [Key - Diagnosis/Classification Variable: Normal:Clin = Normal Clinical Diagnosis, Normal:Alg = Normal Algorithmic Classification, Normal:Alg-Clin = Normal Algorithmic Classification - Normal Clinical Diagnosis, MCI:Clin = MCI Clinical Diagnosis, MCI:Alg = Normal Algorithmic Classification, MCI:Alg-Clin = MCI Algorithmic Classification - MCI Clinical Diagnosis, Dementia:Clin = Dementia Clinical Diagnosis, Dementia:Alg = Dementia Algorithmic Classification, Dementia:Alg-Clin = Dementia Algorithmic Classification - Dementia Clinical Diagnosis]

| MRI Variable | label | estimate | s.e. | p |
|---|---|---|---|---|
| Union Signature | Dementia:Alg | -0.2513 | 0.0575 | 0.0000 |
| Union Signature | Dementia:Alg-Clin | 0.0007 | 0.0483 | 0.9879 |
| Union Signature | Dementia:Clin | -0.2520 | 0.0581 | 0.0000 |
| Union Signature | MCI:Alg | 0.0929 | 0.0504 | 0.0652 |
| Union Signature | MCI:Alg-Clin | -0.0165 | 0.0397 | 0.6785 |
| Union Signature | MCI:Clin | 0.1094 | 0.0489 | 0.0253 |
| Union Signature | Normal:Alg | 0.3242 | 0.0420 | 0.0000 |
| Union Signature | Normal:Alg-Clin | 0.0056 | 0.0261 | 0.8314 |
| Union Signature | Normal:Clin | 0.3186 | 0.0420 | 0.0000 |
| Free Water Cingulum-Hippocampus | Dementia:Alg | 0.2841 | 0.0595 | 0.0000 |
| Free Water Cingulum-Hippocampus | Dementia:Alg-Clin | -0.0125 | 0.0516 | 0.8086 |
| Free Water Cingulum-Hippocampus | Dementia:Clin | 0.2966 | 0.0600 | 0.0000 |
| Free Water Cingulum-Hippocampus | MCI:Alg | -0.0998 | 0.0516 | 0.0529 |
| Free Water Cingulum-Hippocampus | MCI:Alg-Clin | 0.0128 | 0.0430 | 0.7664 |
| Free Water Cingulum-Hippocampus | MCI:Clin | -0.1126 | 0.0494 | 0.0226 |

| | | | | |
|---|---|---|---|---|
| Free Water Cingulum-Hippocampus | Normal:Alg | -0.3435 | 0.0405 | 0.0000 |
| Free Water Cingulum-Hippocampus | Normal:Alg-Clin | 0.0041 | 0.0277 | 0.8834 |
| Free Water Cingulum-Hippocampus | Normal:Clin | -0.3475 | 0.0406 | 0.0000 |
| Hippocampus | Dementia:Alg | -0.2342 | 0.0566 | 0.0000 |
| Hippocampus | Dementia:Alg-Clin | 0.0031 | 0.0438 | 0.9434 |
| Hippocampus | Dementia:Clin | -0.2373 | 0.0571 | 0.0000 |
| Hippocampus | MCI:Alg | 0.0382 | 0.0505 | 0.4494 |
| Hippocampus | MCI:Alg-Clin | -0.0073 | 0.0360 | 0.8397 |
| Hippocampus | MCI:Clin | 0.0455 | 0.0492 | 0.3557 |
| Hippocampus | Normal:Alg | 0.2565 | 0.0436 | 0.0000 |
| Hippocampus | Normal:Alg-Clin | 0.0013 | 0.0237 | 0.9563 |
| Hippocampus | Normal:Clin | 0.2552 | 0.0435 | 0.0000 |
| Cerebral Gray Matter | Dementia:Alg | -0.3141 | 0.0626 | 0.0000 |
| Cerebral Gray Matter | Dementia:Alg-Clin | -0.0253 | 0.0591 | 0.6679 |
| Cerebral Gray Matter | Dementia:Clin | -0.2887 | 0.0634 | 0.0000 |
| Cerebral Gray Matter | MCI:Alg | 0.0792 | 0.0536 | 0.1392 |
| Cerebral Gray Matter | MCI:Alg-Clin | -0.0043 | 0.0485 | 0.9297 |
| Cerebral Gray Matter | MCI:Clin | 0.0835 | 0.0516 | 0.1057 |
| Cerebral Gray Matter | Normal:Alg | 0.3076 | 0.0420 | 0.0000 |
| Cerebral Gray Matter | Normal:Alg-Clin | 0.0062 | 0.0319 | 0.8455 |
| Cerebral Gray Matter | Normal:Clin | 0.3013 | 0.0420 | 0.0000 |

Supplemental Table 6. Model performance comparison for longitudinal imaging measure associations with baseline diagnosis/classification variables. [Key - Diagnosis/Classification Variable: Clinical Diagnosis = Clinical categorical diagnosis, Algorithmic Classification = Algorithmic categorical classification, prob(Normal) = Algorithmically predicted probability of a Normal clinical diagnosis, prob(MCI) = Algorithmically predicted probability of a MCI clinical diagnosis, prob(Dementia) = Algorithmically predicted probability of a Dementia clinical diagnosis; AIC = Akaike Information Criterion, BIC = Bayesian Information Criterion, R2 = R-Squared, RMSE = Root Mean Square Error]

| MRI Variable | Diagnosis/Classification Variable | AIC | BIC | R2_marginal |
|---|---|---|---|---|
| Union Signature | Clinical Diagnosis | 3014 | 3056 | 0.256 |
| Union Signature | Algorithmic Classification | 3188 | 3230 | 0.244 |
| Union Signature | prob(Normal) | 3121 | 3152 | 0.304 |
| Union Signature | prob(MCI) | 3179 | 3210 | 0.247 |
| Union Signature | prob(Dementia) | 3198 | 3229 | 0.209 |
| Freewater Cingulum-Hippocampus | Clinical Diagnosis | 2558 | 2598 | 0.187 |
| Freewater Cingulum-Hippocampus | Algorithmic Classification | 2628 | 2669 | 0.2 |
| Freewater Cingulum-Hippocampus | prob(Normal) | 2587 | 2617 | 0.248 |
| Freewater Cingulum-Hippocampus | prob(MCI) | 2626 | 2656 | 0.207 |
| Freewater Cingulum-Hippocampus | prob(Dementia) | 2632 | 2662 | 0.187 |
| Hippocampus | Clinical Diagnosis | 2914 | 2956 | 0.253 |
| Hippocampus | Algorithmic Classification | 3012 | 3054 | 0.234 |
| Hippocampus | prob(Normal) | 2982 | 3014 | 0.261 |
| Hippocampus | prob(MCI) | 3038 | 3070 | 0.197 |

| | | | | |
|---|---|---|---|---|
| Hippocampus | prob(Dementia) | 3036 | 3067 | 0.184 |
| Cerebral Gray Matter | Clinical Diagnosis | 3335 | 3377 | 0.172 |
| Cerebral Gray Matter | Algorithmic Classification | 3466 | 3508 | 0.167 |
| Cerebral Gray Matter | prob(Normal) | 3421 | 3453 | 0.206 |
| Cerebral Gray Matter | prob(MCI) | 3453 | 3485 | 0.176 |
| Cerebral Gray Matter | prob(Dementia) | 3481 | 3513 | 0.133 |

Supplemental Table 7. Baseline diagnosis/classification effects by diagnosis type on brain measure intercept in longitudinal models (ADRC cohort). (Time-from-1st-scan and baseline diagnosis/classification are fixed effects independent variables in mixed effects longitudinal models of longitudinal change in brain measures. p values are not corrected for multiple comparisons.)[Key: Normal:Clin = Normal Clinical Diagnosis, Normal:Alg = Normal Algorithmic Classification, Normal:Alg-Clin = Normal Algorithmic Classification - Normal Clinical Diagnosis, MCI:Clin = MCI Clinical Diagnosis, MCI:Alg = Normal Algorithmic Classification, MCI:Alg-Clin = MCI Algorithmic Classification - MCI Clinical Diagnosis, Dementia:Clin = Dementia Clinical Diagnosis, Dementia:Alg = Dementia Algorithmic Classification, Dementia:Alg-Clin = Dementia Algorithmic Classification - Dementia Clinical Diagnosis]

| MRI Variable | label | estimate | s.e. | p |
|---|---|---|---|---|
| Union Signature | Normal:Clin | 0.3191 | 0.0457 | 0.0000 |
| Union Signature | Normal:Alg | 0.3109 | 0.0453 | 0.0000 |
| Union Signature | Normal:Alg-Clin | -0.0082 | 0.0281 | 0.7705 |
| Union Signature | MCI:Clin | 0.0303 | 0.0596 | 0.6108 |
| Union Signature | MCI:Alg | 0.0557 | 0.0617 | 0.3660 |
| Union Signature | MCI:Alg-Clin | 0.0254 | 0.0467 | 0.5864 |
| Union Signature | Dementia:Clin | -0.0404 | 0.1267 | 0.7501 |
| Union Signature | Dementia:Alg | -0.0781 | 0.1156 | 0.4989 |
| Union Signature | Dementia:Alg-Clin | -0.0378 | 0.1144 | 0.7413 |
| Free Water Cingulum-Hippocampus | Normal:Clin | -0.3372 | 0.0450 | 0.0000 |
| Free Water Cingulum-Hippocampus | Normal:Alg | -0.3320 | 0.0445 | 0.0000 |
| Free Water Cingulum-Hippocampus | Normal:Alg-Clin | 0.0051 | 0.0304 | 0.8657 |
| Free Water Cingulum-Hippocampus | MCI:Clin | -0.0587 | 0.0630 | 0.3519 |
| Free Water Cingulum-Hippocampus | MCI:Alg | -0.0531 | 0.0664 | 0.4235 |
| Free Water Cingulum-Hippocampus | MCI:Alg-Clin | 0.0055 | 0.0532 | 0.9171 |
| Free Water Cingulum-Hippocampus | Dementia:Clin | -0.0198 | 0.1352 | 0.8838 |
| Free Water Cingulum-Hippocampus | Dementia:Alg | -0.0442 | 0.1252 | 0.7240 |

| | | | | |
|---|---|---|---|---|
| Free Water Cingulum-Hippocampus | Dementia:Alg-Clin | -0.0245 | 0.1308 | 0.8517 |
| Hippocampus | Normal:Clin | 0.2393 | 0.0469 | 0.0000 |
| Hippocampus | Normal:Alg | 0.2371 | 0.0466 | 0.0000 |
| Hippocampus | Normal:Alg-Clin | -0.0022 | 0.0257 | 0.9330 |
| Hippocampus | MCI:Clin | 0.0331 | 0.0590 | 0.5746 |
| Hippocampus | MCI:Alg | 0.0441 | 0.0608 | 0.4682 |
| Hippocampus | MCI:Alg-Clin | 0.0110 | 0.0426 | 0.7967 |
| Hippocampus | Dementia:Clin | -0.1403 | 0.1203 | 0.2434 |
| Hippocampus | Dementia:Alg | -0.1373 | 0.1100 | 0.2120 |
| Hippocampus | Dementia:Alg-Clin | 0.0030 | 0.1045 | 0.9772 |
| Cerebral Gray Matter | Normal:Clin | 0.3171 | 0.0459 | 0.0000 |
| Cerebral Gray Matter | Normal:Alg | 0.3132 | 0.0454 | 0.0000 |
| Cerebral Gray Matter | Normal:Alg-Clin | -0.0038 | 0.0344 | 0.9109 |
| Cerebral Gray Matter | MCI:Clin | -0.0355 | 0.0637 | 0.5774 |
| Cerebral Gray Matter | MCI:Alg | -0.0356 | 0.0663 | 0.5911 |
| Cerebral Gray Matter | MCI:Alg-Clin | -0.0001 | 0.0569 | 0.9983 |
| Cerebral Gray Matter | Dementia:Clin | -0.0087 | 0.1437 | 0.9516 |
| Cerebral Gray Matter | Dementia:Alg | -0.1131 | 0.1305 | 0.3860 |
| Cerebral Gray Matter | Dementia:Alg-Clin | -0.1044 | 0.1393 | 0.4535 |

Supplemental Table 8. Time-by-baseline diagnosis/classification effects by diagnosis/classification type on brain measure in longitudinal models (ADRC cohort). Results show average annual rate of change in brain measure associated with diagnosis/classification variables and difference in average rate of change across diagnosis/classification types. (Time-from-1st-scan and baseline diagnosis/classification are fixed effects independent variables in mixed effects longitudinal models of longitudinal change in brain measures. p values are not corrected for multiple comparisons.) [Key: Normal:Clin = Normal Clinical Diagnosis, Normal:Alg = Normal Algorithmic Classification, Normal:Alg-Clin = Normal Algorithmic Classification - Normal Clinical Diagnosis, MCI:Clin = MCI Clinical Diagnosis, MCI:Alg = Normal Algorithmic Classification, MCI:Alg-Clin = MCI Algorithmic Classification - MCI Clinical Diagnosis, Dementia:Clin = Dementia Clinical Diagnosis, Dementia:Alg = Dementia Algorithmic Classification, Dementia:Alg-Clin = Dementia Algorithmic Classification - Dementia Clinical Diagnosis]

| MRI Variable | label | estimate | s.e. | p |
|---|---|---|---|---|
| Union Signature | Normal:Clin | -0.0944 | 0.0046 | 0.0000 |
| Union Signature | Normal:Alg | -0.0919 | 0.0044 | 0.0000 |
| Union Signature | Normal:Alg-Clin | 0.0025 | 0.0057 | 0.6616 |
| Union Signature | MCI:Clin | -0.0989 | 0.0088 | 0.0000 |
| Union Signature | MCI:Alg | -0.1128 | 0.0096 | 0.0000 |
| Union Signature | MCI:Alg-Clin | -0.0138 | 0.0119 | 0.2461 |
| Union Signature | Dementia:Clin | -0.2136 | 0.0279 | 0.0000 |
| Union Signature | Dementia:Alg | -0.1731 | 0.0249 | 0.0000 |
| Union Signature | Dementia:Alg-Clin | 0.0405 | 0.0355 | 0.2544 |
| Free Water Cingulum-Hippocampus | Normal:Clin | 0.0920 | 0.0051 | 0.0000 |
| Free Water Cingulum-Hippocampus | Normal:Alg | 0.0902 | 0.0050 | 0.0000 |
| Free Water Cingulum-Hippocampus | Normal:Alg-Clin | -0.0018 | 0.0061 | 0.7678 |
| Free Water Cingulum-Hippocampus | MCI:Clin | 0.0940 | 0.0107 | 0.0000 |
| Free Water Cingulum-Hippocampus | MCI:Alg | 0.0933 | 0.0115 | 0.0000 |
| Free Water Cingulum-Hippocampus | MCI:Alg-Clin | -0.0007 | 0.0131 | 0.9567 |

| | | | | |
|---|---|---|---|---|
| Free Water Cingulum-Hippocampus | Dementia:Clin | 0.1772 | 0.0344 | 0.0000 |
| Free Water Cingulum-Hippocampus | Dementia:Alg | 0.1785 | 0.0301 | 0.0000 |
| Free Water Cingulum-Hippocampus | Dementia:Alg-Clin | 0.0013 | 0.0422 | 0.9753 |
| Hippocampus | Normal:Clin | -0.0755 | 0.0042 | 0.0000 |
| Hippocampus | Normal:Alg | -0.0751 | 0.0040 | 0.0000 |
| Hippocampus | Normal:Alg-Clin | 0.0003 | 0.0052 | 0.9524 |
| Hippocampus | MCI:Clin | -0.0923 | 0.0080 | 0.0000 |
| Hippocampus | MCI:Alg | -0.0982 | 0.0088 | 0.0000 |
| Hippocampus | MCI:Alg-Clin | -0.0059 | 0.0109 | 0.5891 |
| Hippocampus | Dementia:Clin | -0.1427 | 0.0255 | 0.0000 |
| Hippocampus | Dementia:Alg | -0.1240 | 0.0228 | 0.0000 |
| Hippocampus | Dementia:Alg-Clin | 0.0187 | 0.0324 | 0.5648 |
| Cerebral Gray Matter | Normal:Clin | -0.0716 | 0.0055 | 0.0000 |
| Cerebral Gray Matter | Normal:Alg | -0.0708 | 0.0054 | 0.0000 |
| Cerebral Gray Matter | Normal:Alg-Clin | 0.0008 | 0.0070 | 0.9072 |
| Cerebral Gray Matter | MCI:Clin | -0.0743 | 0.0106 | 0.0000 |
| Cerebral Gray Matter | MCI:Alg | -0.0775 | 0.0117 | 0.0000 |
| Cerebral Gray Matter | MCI:Alg-Clin | -0.0033 | 0.0146 | 0.8232 |
| Cerebral Gray Matter | Dementia:Clin | -0.2077 | 0.0340 | 0.0000 |
| Cerebral Gray Matter | Dementia:Alg | -0.1393 | 0.0301 | 0.0000 |
| Cerebral Gray Matter | Dementia:Alg-Clin | 0.0685 | 0.0433 | 0.1136 |
